# Impact of COVID-19 on diabetes mellitus outcomes and care in sub-Saharan Africa: A scoping review

**DOI:** 10.1101/2024.04.10.24305598

**Authors:** Wenceslaus Sseguya, Silver Bahendeka, Sara MacLennan, Nimesh Mody, Aravinda Meera Guntupalli

## Abstract

**Background:** The COVID-19 pandemic impacted diabetes mellitus clinical outcomes and chronic care globally. However, little is known about its impact in low-resource settings such as sub-Saharan Africa. Hence, to address this, we systematically conducted a scoping review to explore the COVID-19 impact on diabetes outcomes and care in countries of sub-Saharan Africa.

**Methods:** We applied our search strategy to PubMed, Web of Science, CINAHL, African Index Medicus, Google Scholar, Cochrane Library, Scopus, Science Direct, ERIC and Embase to obtain relevant articles published from January 2020 to March 2023. Two independent reviewers were involved in the screening of retrieved articles. Data from eligible articles were extracted from quantitative, qualitative and mixed methods studies. Numerical data were summarised using descriptive statistics, while a thematic framework was used to categorise and identify themes for qualitative data.

**Results:** We found 42 of the retrieved 360 articles eligible, mainly from South Africa, Ethiopia and Ghana (73.4%). COVID-19 increased the risk of death (OR 1.30 – 9.0, 95% CI), hospitalisation (OR 3.30 – 3.73: 95% CI), and severity (OR: 1.30-4.05, 95% CI) in persons with diabetes mellitus. COVID-19 also increased the risk of developing diabetes mellitus in hospitalised cases. The pandemic, on the other hand, was associated with disruptions in patient self-management routine and diabetes mellitus care service delivery. Three major themes emerged, namely, (i) patient-related health management challenges, (ii) diabetes mellitus care service delivery challenges, and (iii) reorganisation of diabetes mellitus care delivery.

**Conclusion:** COVID-19 increased mortality and morbidity among people living with diabetes mellitus. In addition, the COVID-19 pandemic worsened diabetes mellitus care management. Sub-Saharan African countries should, therefore, institute appropriate policy considerations for persons with diabetes mellitus during widespread emergencies.

## Introduction

Global evidence suggests that the coronavirus disease 2019 (COVID-19) resulted in a worldwide surge in mortality, morbidity, and disability, which predominantly occurred among older adults and individuals with chronic disease conditions [1,2]. COVID-19 has been reported to worsen diabetes mellitus (DM) clinical outcomes in particular, and DM care in general generally [3–8]. However, very little in this context is known in low- and middle-income countries, particularly in sub-Saharan Africa (SSA).

While SSA is estimated to be host to 24 million of the estimated 537 million people with DM globally, the region records the highest rate of DM-related premature mortality [9]. Furthermore, SSA is predicted to experience the highest rate of rise in DM prevalence than any other region by 2040, depicting the magnitude of a growing threat [9]. DM is an under-researched area in SSA, which may underlie the limited understanding of the scale of the COVID-19 impact on persons living with DM (PLWD) and related vulnerabilities within the region. To address this gap, we carried out a scoping review to assimilate knowledge in this area that supports evidence-based policy consideration and stimulates future research in this field in SSA.

We, therefore systematically conducted a scoping review of published qualitative, quantitative and mixed methods literature to explore the COVID-19 impact on DM outcomes and care in SSA. Our scoping review aimed to: (i) identify and characterise impact of COVID-19 infection on clinical outcomes of DM; (ii) describe DM care aspects that were impacted by the COVID-19 pandemic; and (iii) identify existing gaps in knowledge and research.

## Methods

### Study design

We report our scoping review in line with the Preferred Reporting Items for Systematic Reviews and Meta-analyses extension for Scoping Reviews (PRISMA-ScR) (S1 PRISMA-ScR Checklist). The initial protocol for this scoping review is reposited with Open Science Framework [https://doi.org/10.17605/OSF.IO/9JCKF].

### Data sources and search strategy

We searched ten electronic databases, i.e., PubMed, Web of Science, Cumulative Index to Nursing and Allied Health Literature (CINAHL), African Index Medicus, Google Scholar, Cochrane Library, Scopus, Science Direct, Education Resource Information Centre (ERIC) and Embase. We defined our search strategy guided by the SPIDER (Sample population, Phenomenon of Interest, Design, Evaluation and Research type) framework as outlined by Cooke et al. [10] to identify relevant literature from qualitative and mixed methods studies. Additionally, to capture relevant literature from quantitative studies, we enriched our search strategy by incorporating appropriate elements of the PICO (Population, Intervention, Comparison and Outcome) framework [11]. The detailed search strategy applied to all citation databases with their respective search strings is provided as supplementary material (S2 Search strategy). A search across all databases was initially conducted in May 2022 and later updated using the same search strategy in March 2023 to include any relevant records published between the two periods. This also opened up possibilities for including studies with data on various ‘waves’ of COVID-19 infection and emerging interventions as the pandemic progressed. All retrieved records were merged into a single MS^®^ Excel file for subsequent removal of duplicate records and screening.

### Selection criteria

The retrieved records were screened for eligibility through two stages, i.e., an initial review of article title and abstract and a subsequent full-text review of articles to be considered in final inclusion. An initial screening for the title and abstract was independently conducted by WS and AMG and reviewed by SB, who also resolved any disagreements in screening decisions. The same approach was applied for full-text screening. We defined agreement as a matching decision independently held by the reviewers involved in the screening process.

The inclusion criteria were (i) articles from any country listed under SSA by the World Bank in 2021 [12], (ii) articles focusing on or concerning DM and COVID-19, (ii) peer-reviewed articles and reports and (iv) published from 01 January 2020 – 22 March 2023. The exclusion criteria were (i) no full-text availability, (ii) articles not published in the English language, (iii) non-human studies, (iv) reviews, (v) articles with irrelevant scope, (vi) duplicate articles, and (vii) articles published as multicountry studies involving countries outside SSA but without disaggregation of country-specific data (Fig 1).

**Fig. 1:**
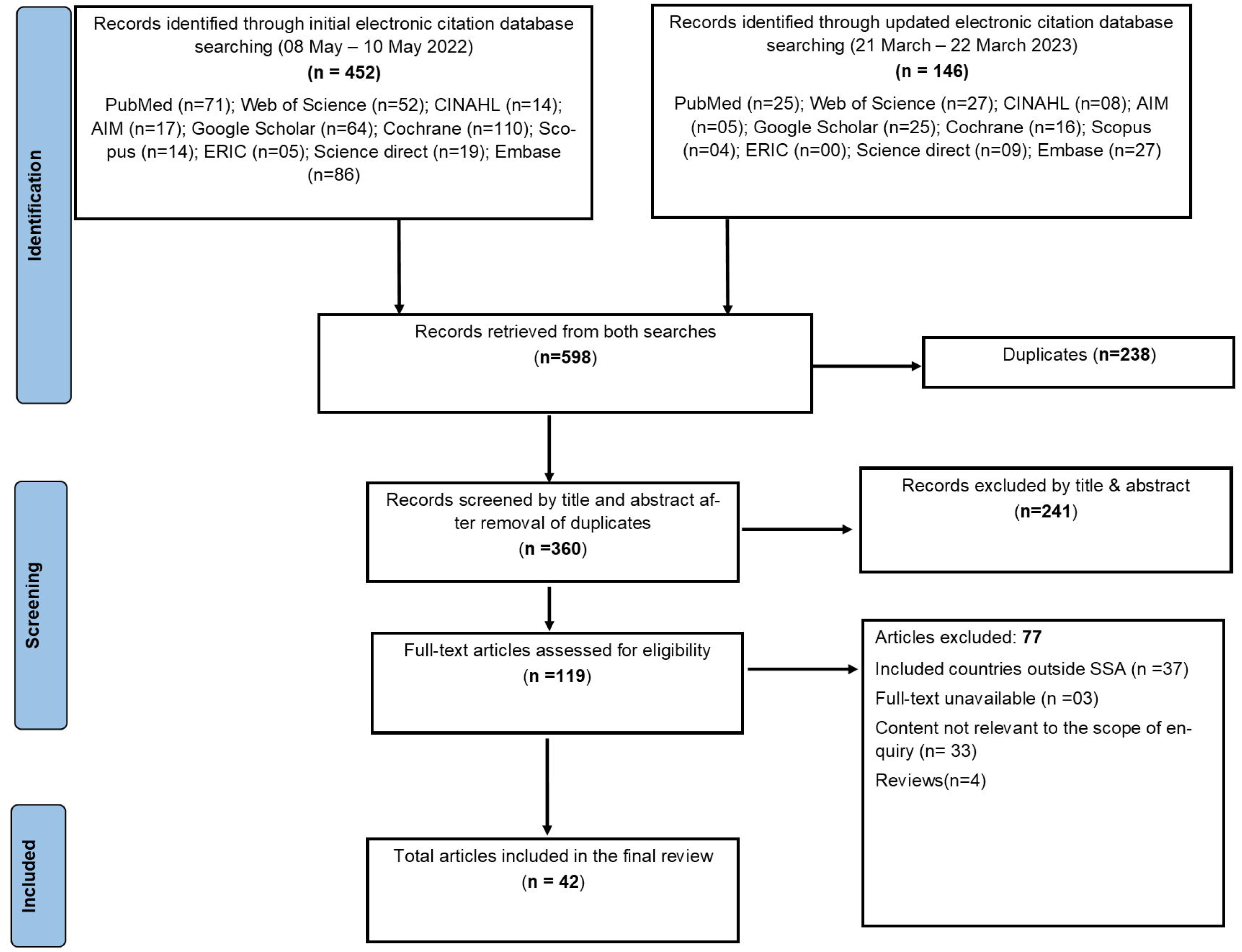
PRISMA-ScR diagram reporting outcomes of the systematic scoping review process.

### Data extraction and management

Data variables of interest from the selected articles were extracted and charted in the extraction form. The data extraction form was developed by WS and reviewed by AMG, SB, and SM. It was then tested with two randomly selected articles from each set of quantitative, qualitative, and mixed methods studies for appropriateness. Appropriate revisions were made and continuously refined and updated throughout the data extraction process. Data extraction and charting were conducted by WS and independently reviewed by AMG and SB during the extraction and charting phase.

### Data synthesis

We used an inductive thematic approach to synthesise and collate findings of qualitative and mixed-methods studies and open-ended results of quantitative studies.

We used SPSS^®^ version 27.0 (IBM Corp, Armonk: New York) to summarise findings from quantitative studies as mean (SD), range (minimum and maximum), proportions and frequencies, where appropriate. Due to the variability in methodological designs of interventions and outcome measures across studies, a meta-analysis was not performed.

## Results

### Selection and characteristics of included studies

A total of 360 unique records were retrieved from database searches, 42 of which were eligible for final inclusion (Fig 1). Inter-reviewer reliability analysis using the Cohen’s kappa showed substantial agreement between reviewers at title and abstract screening (k=0.626, p<0.01), and moderate agreement at full-text screening (k=0.545, p<0.01). The detailed description of information of the included studies is shown in Table 1.

**Table 1:**
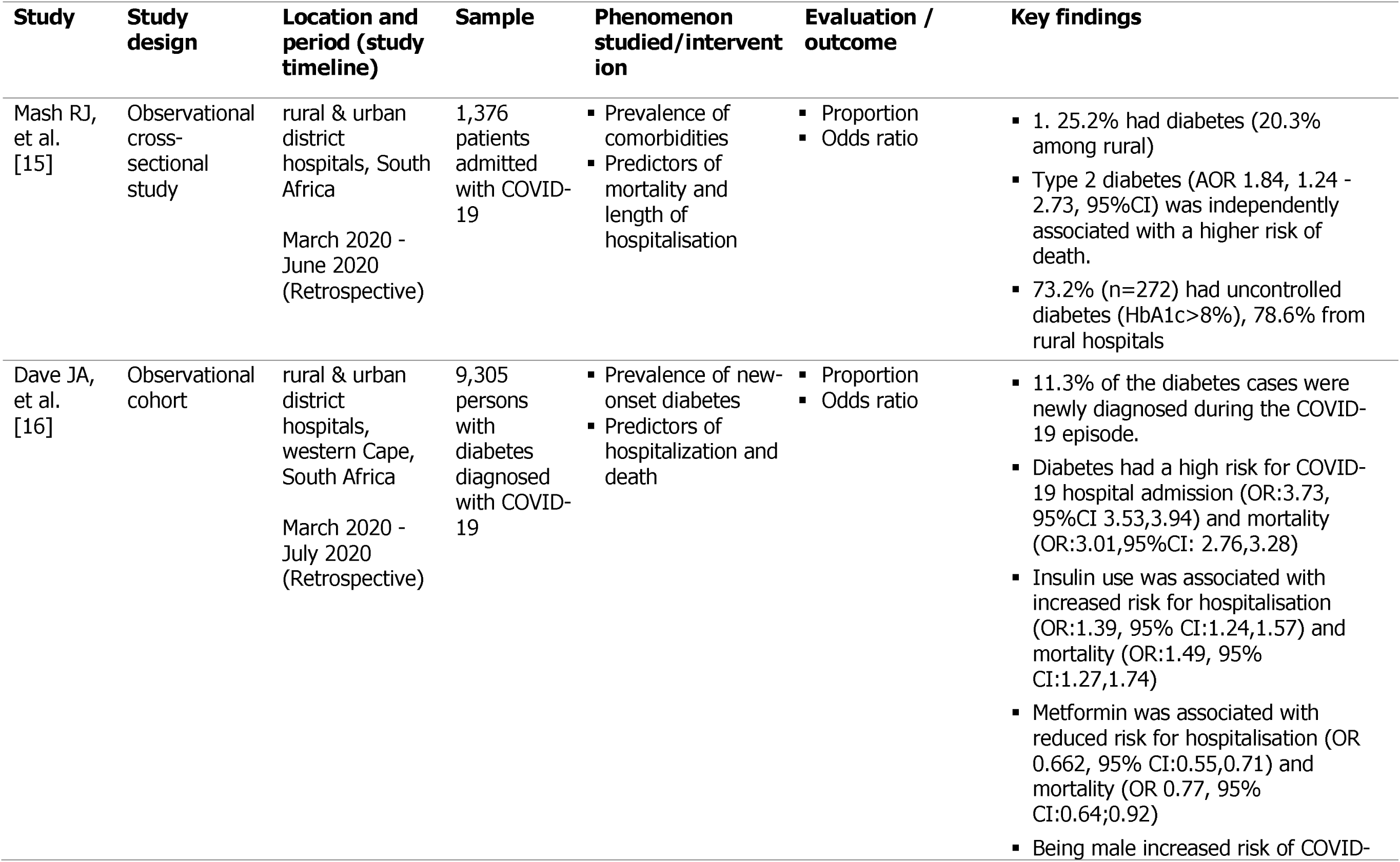
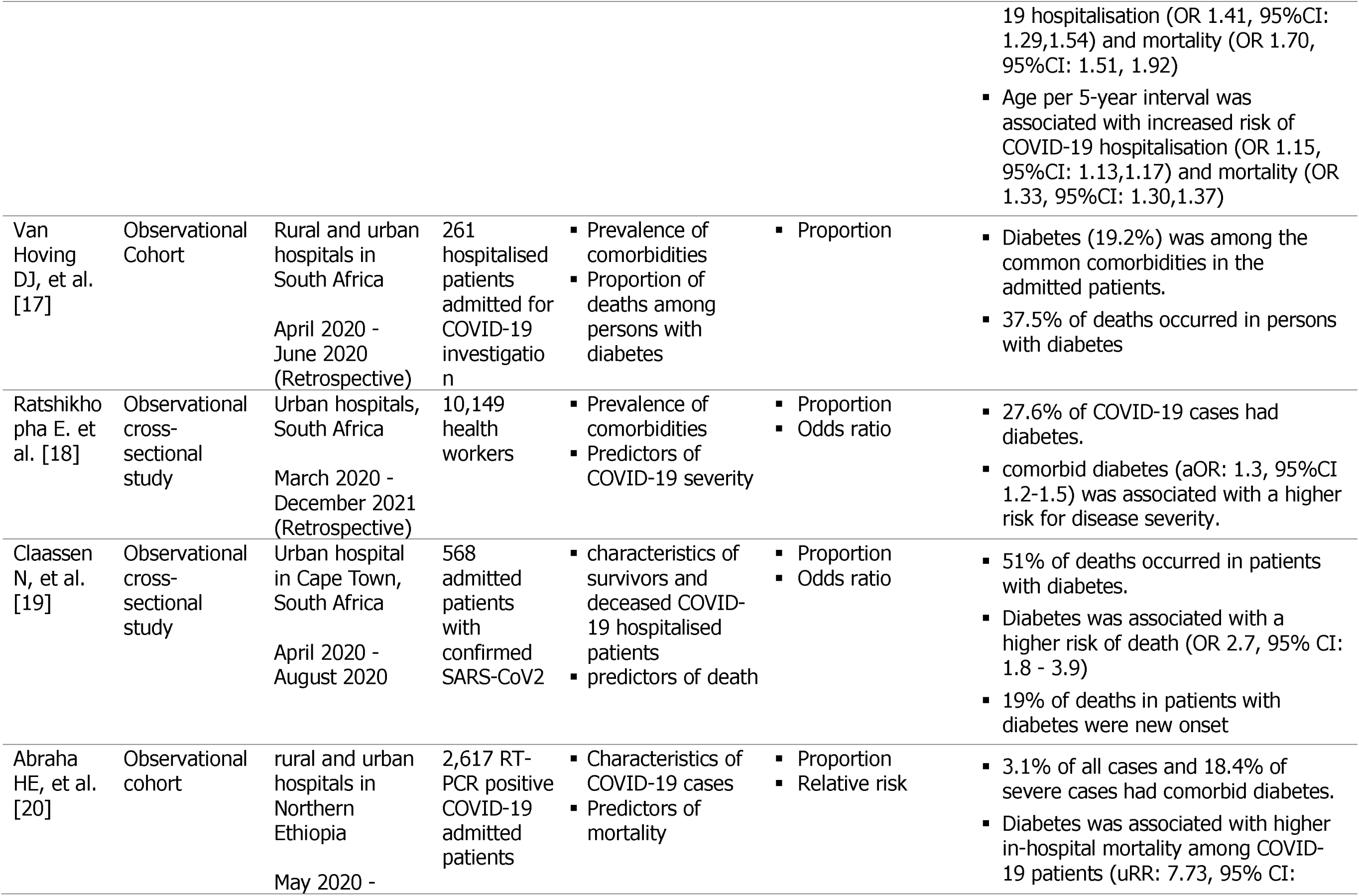
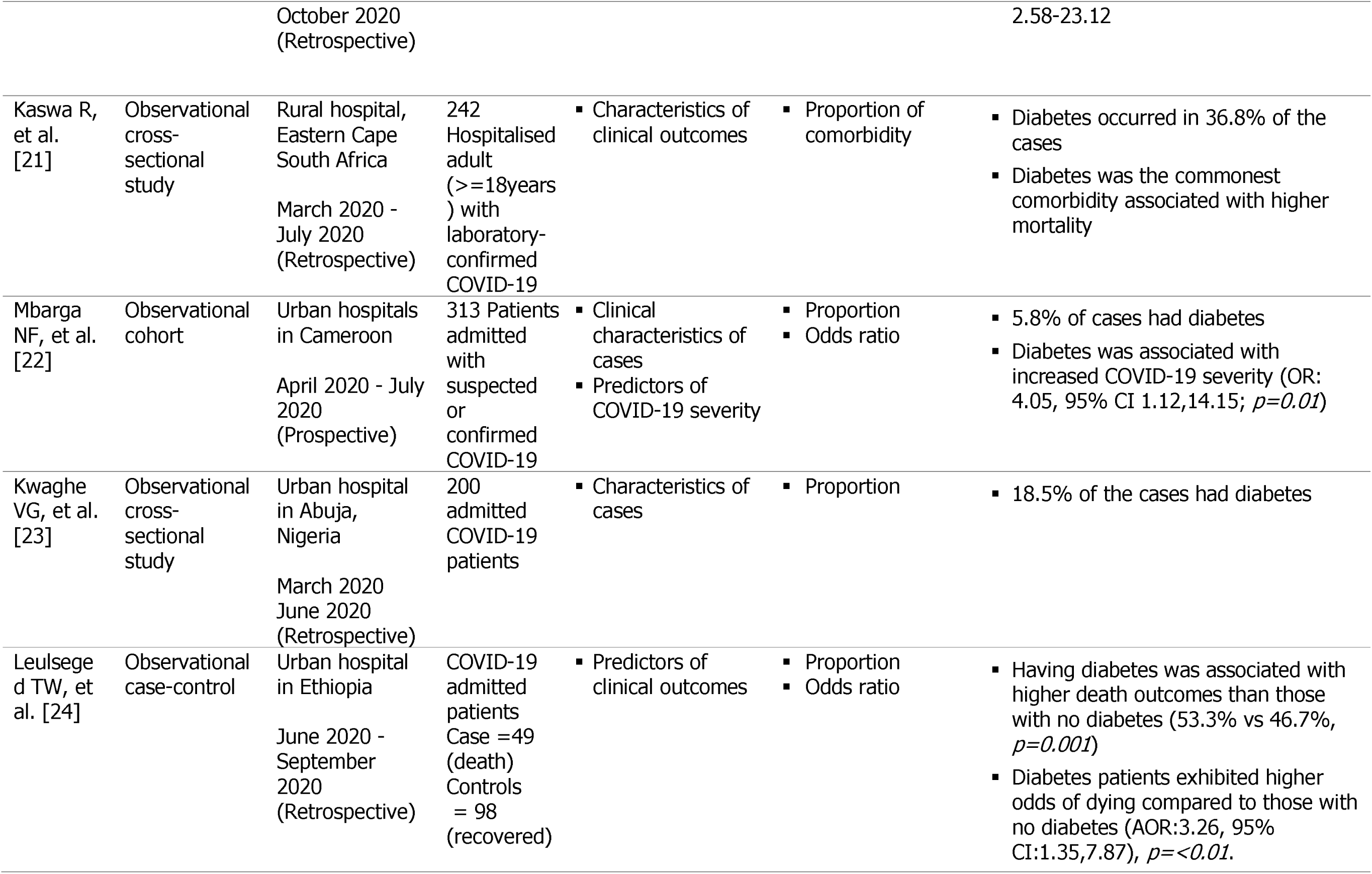
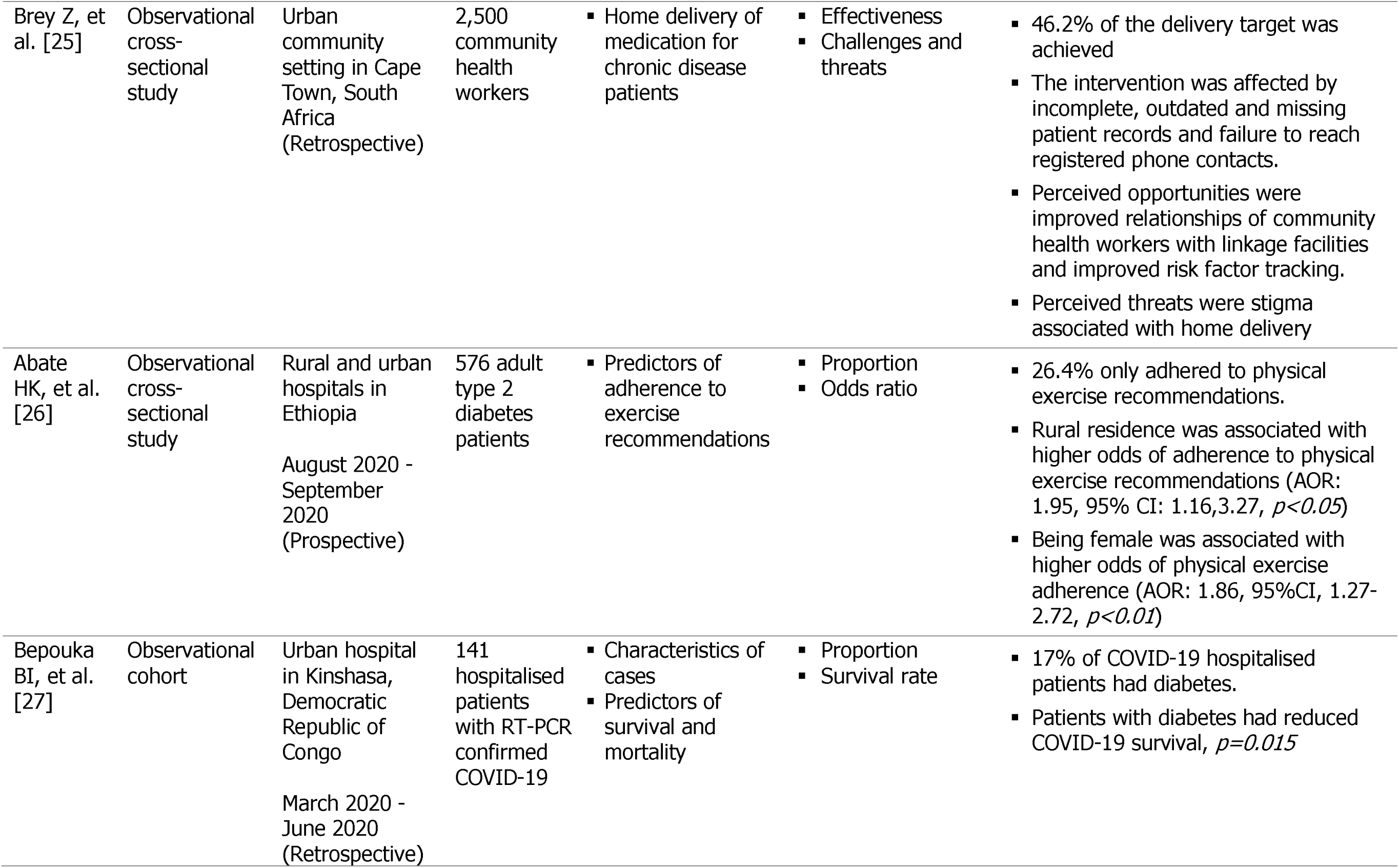
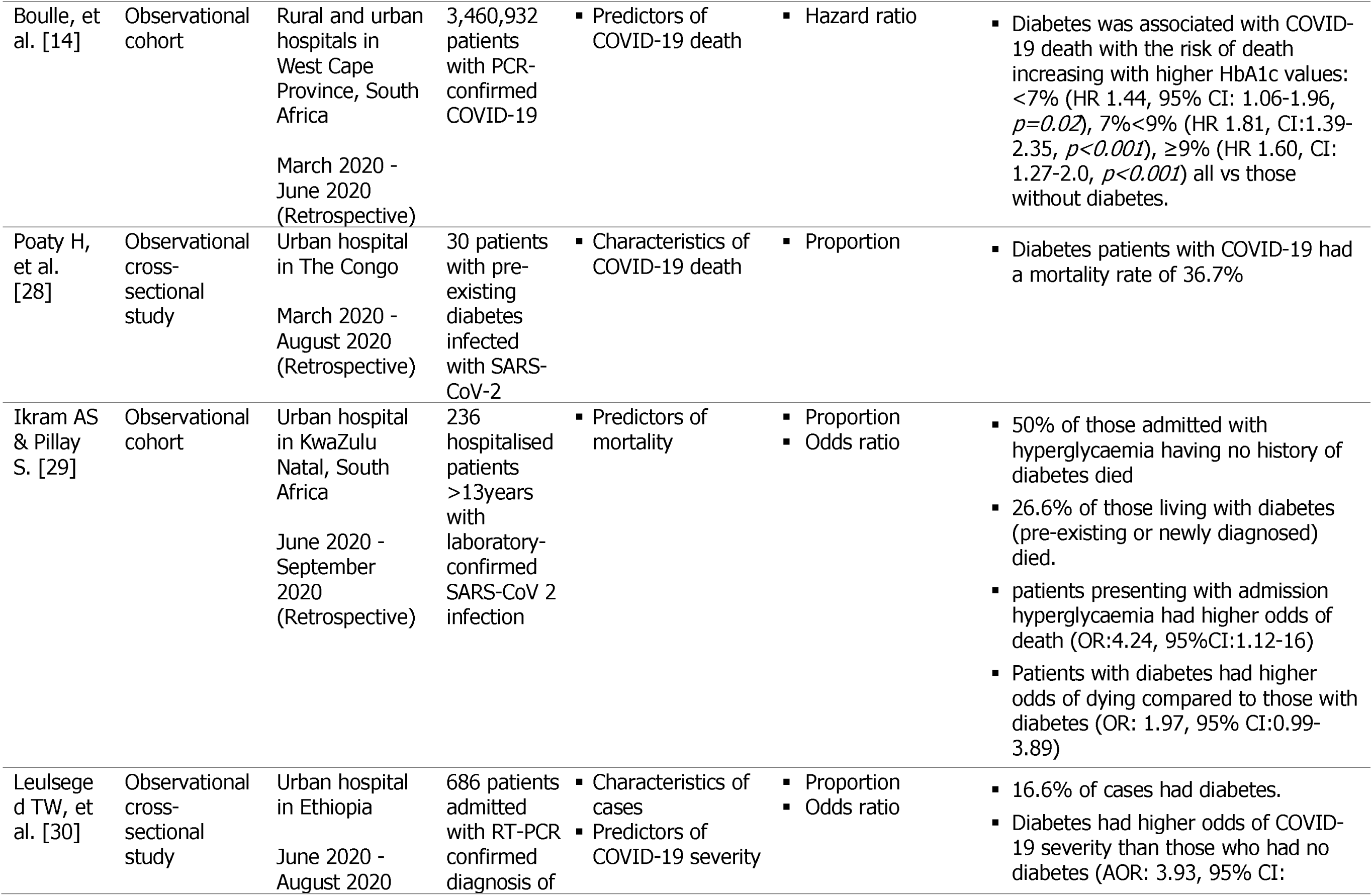
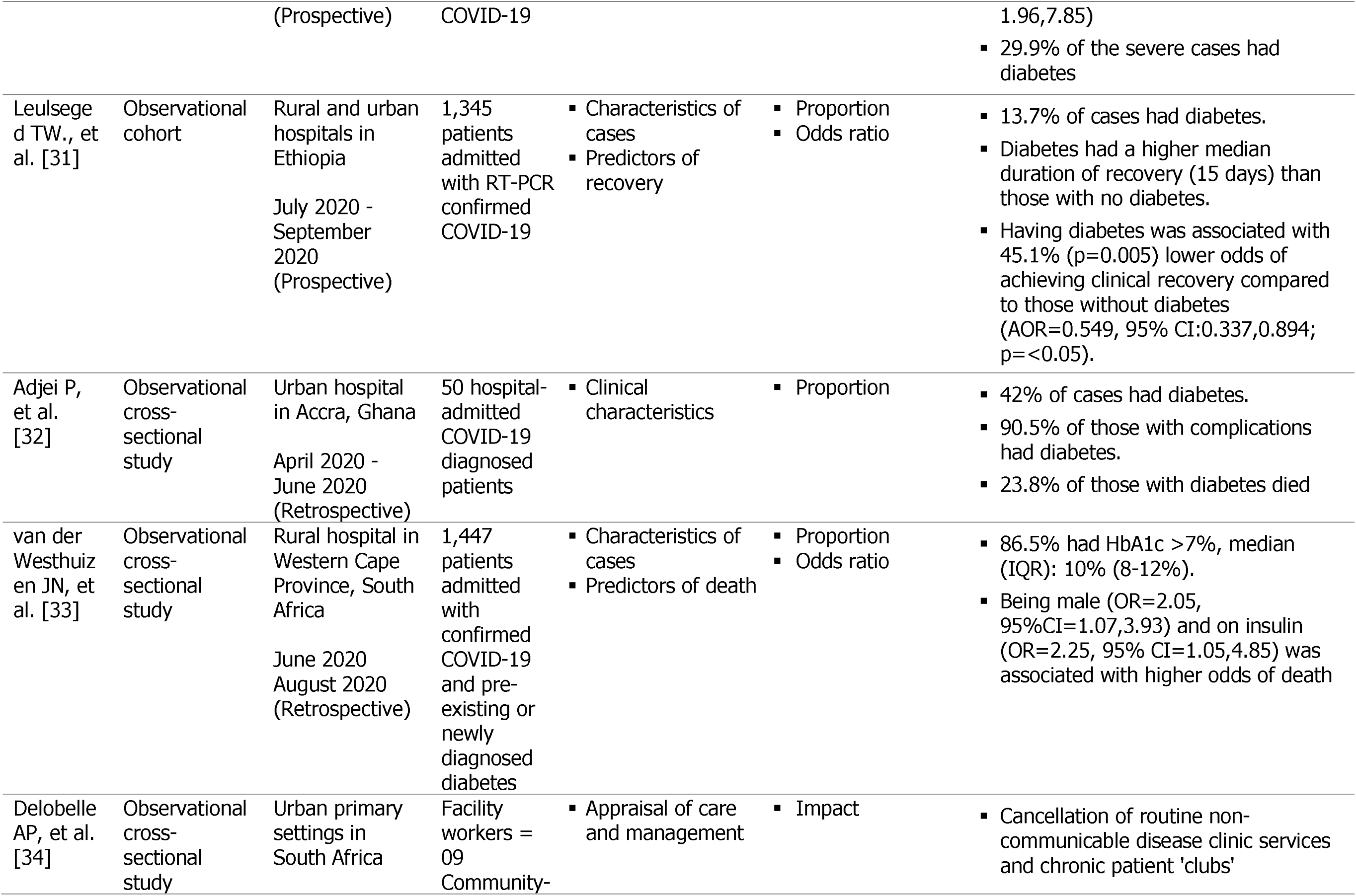
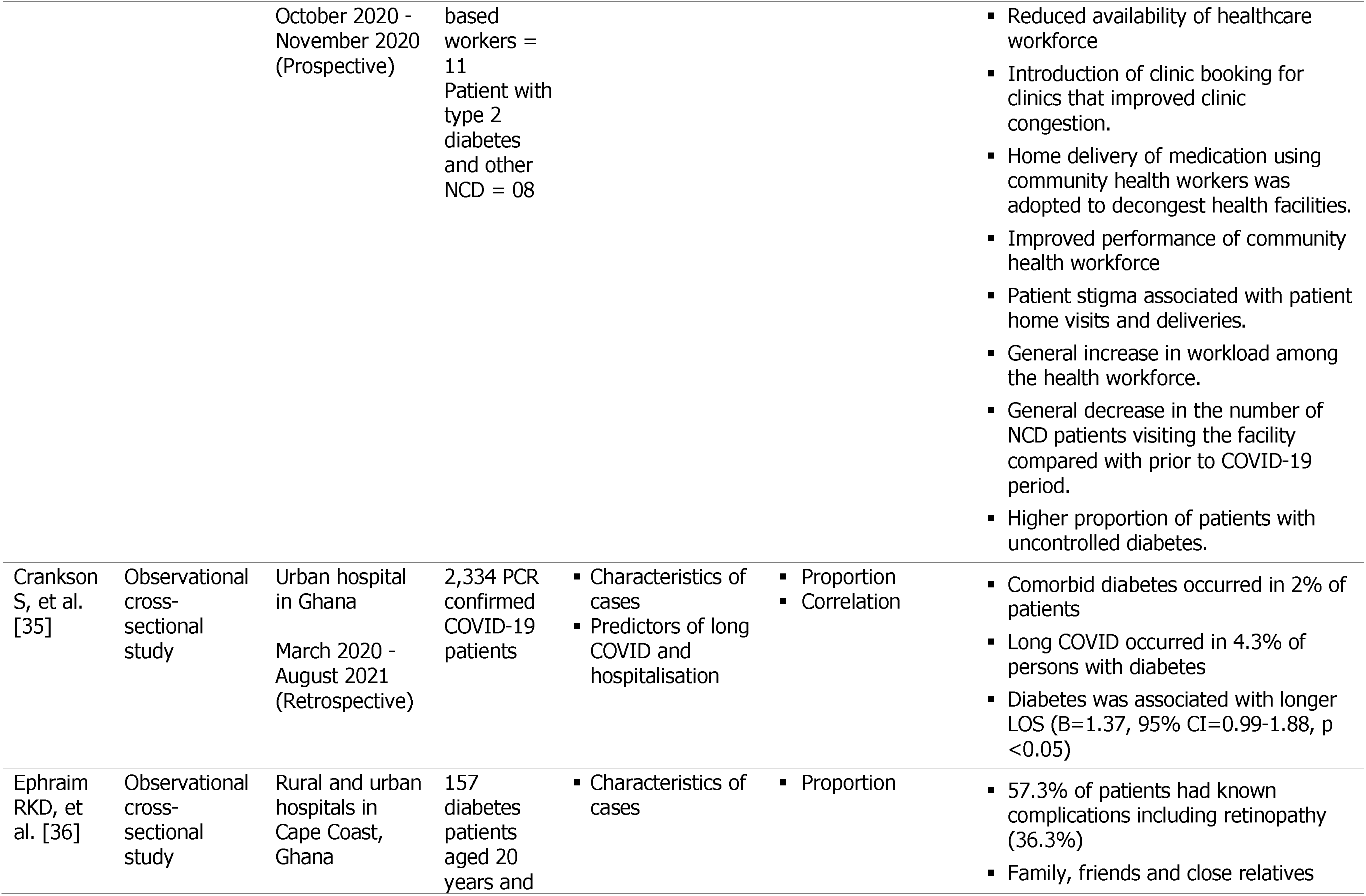
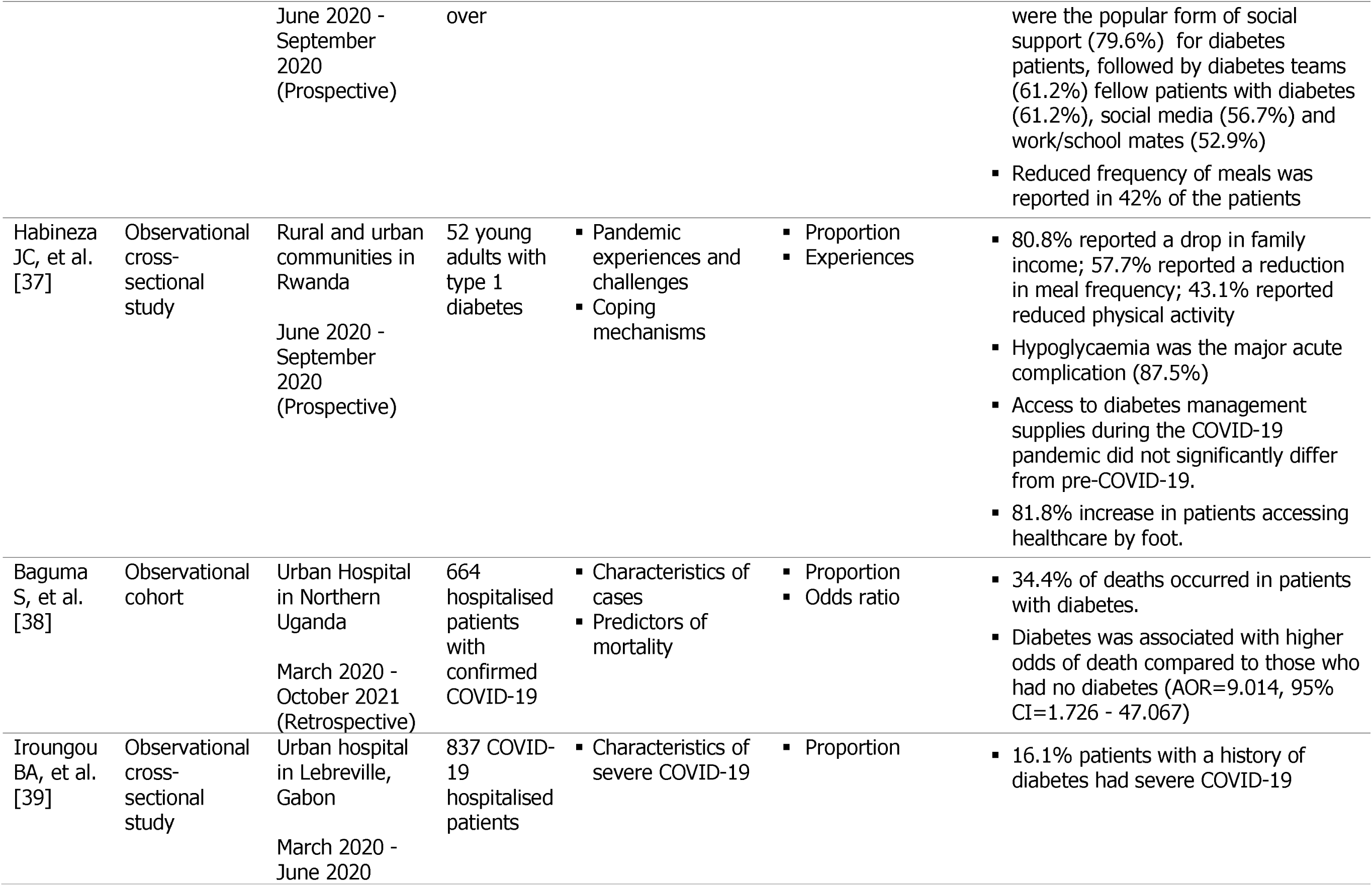
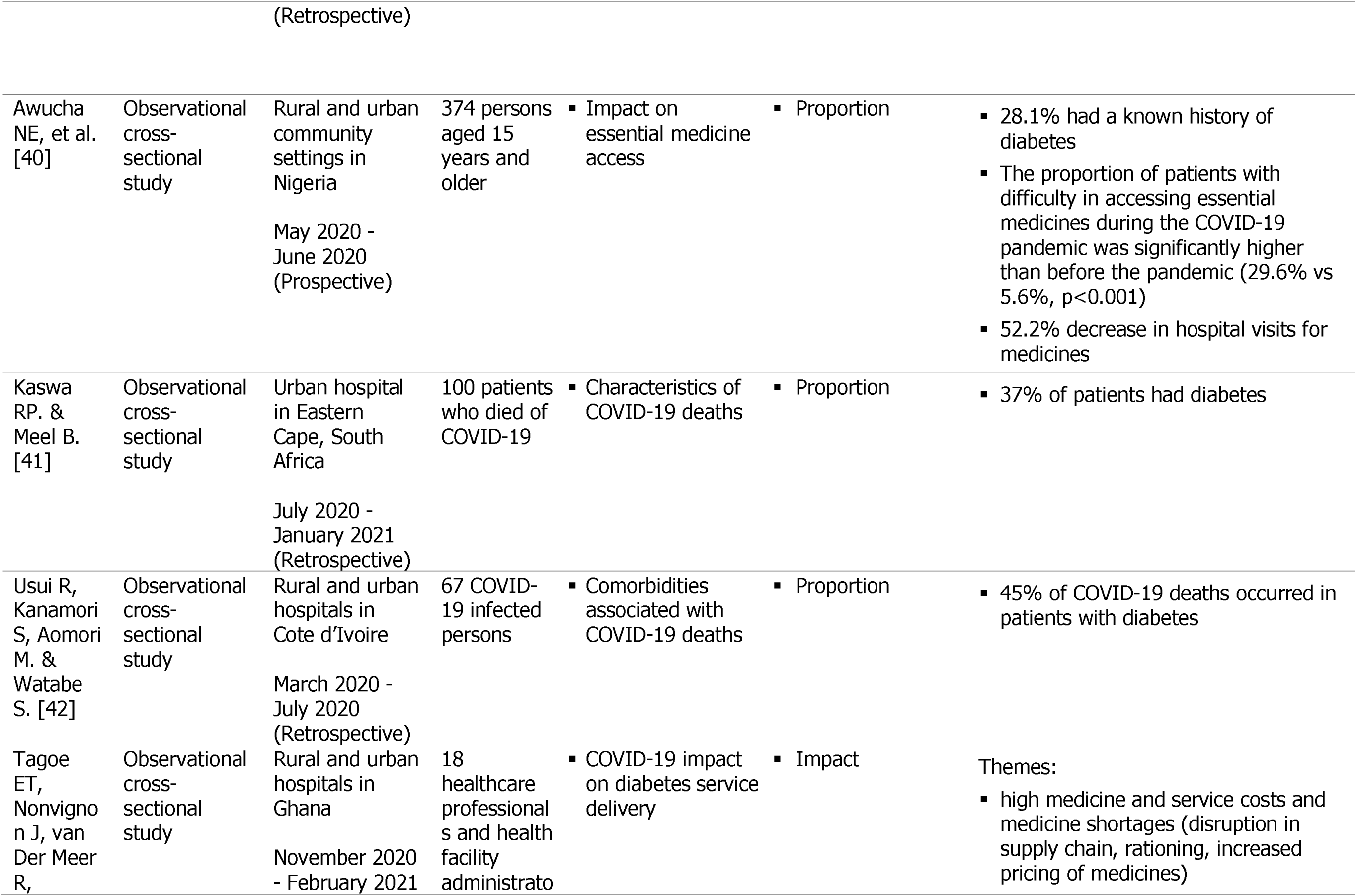
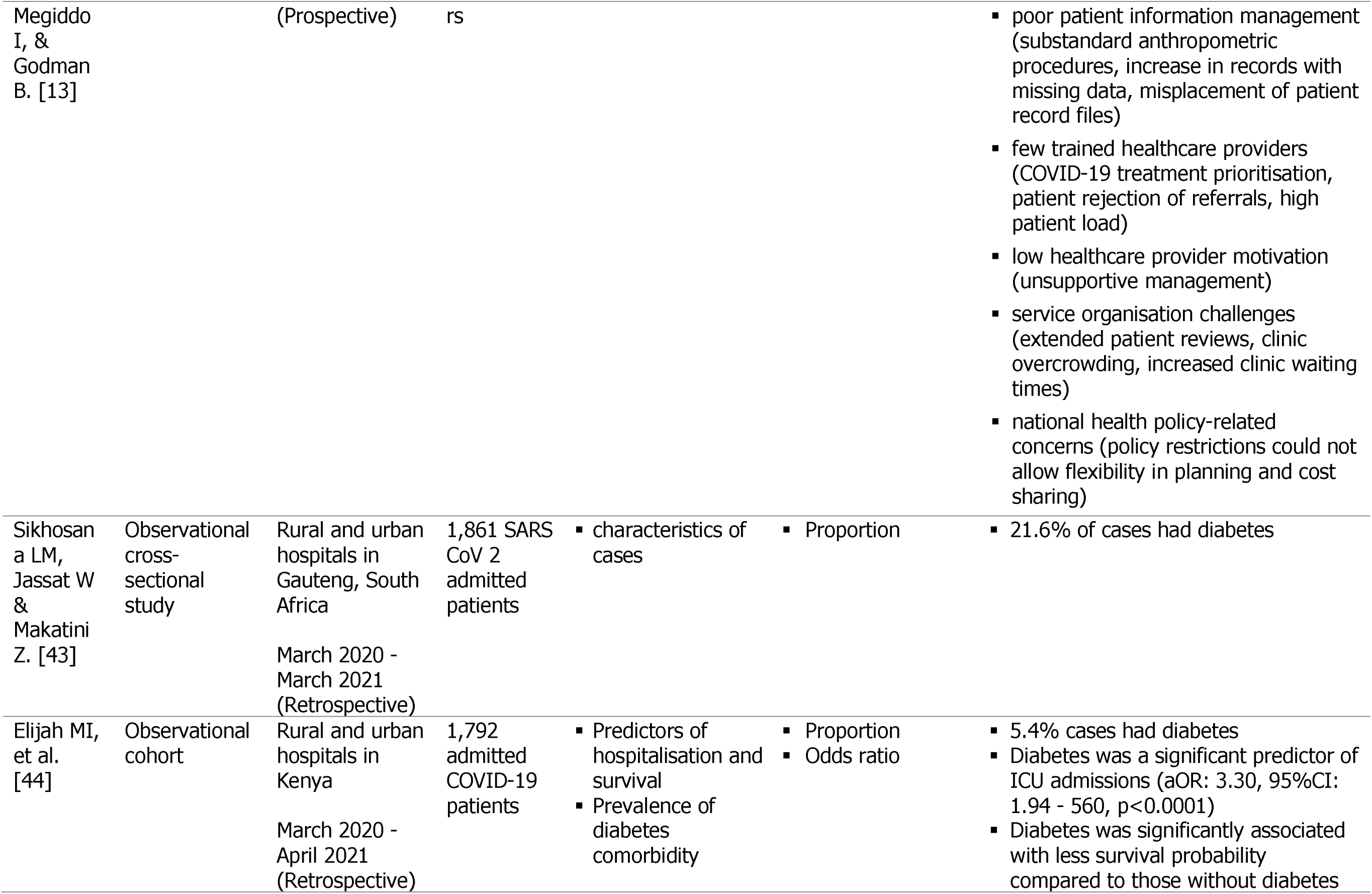
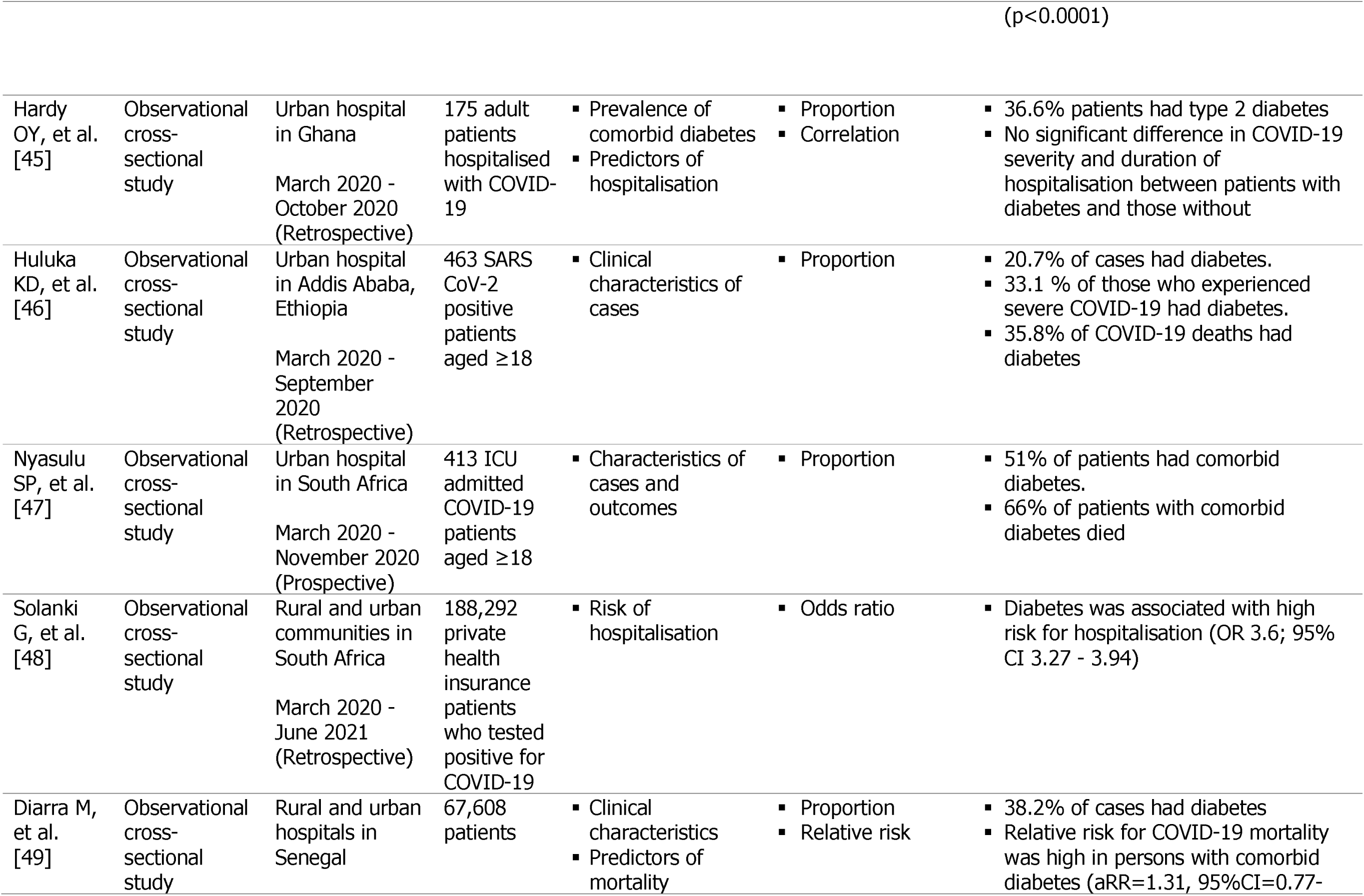
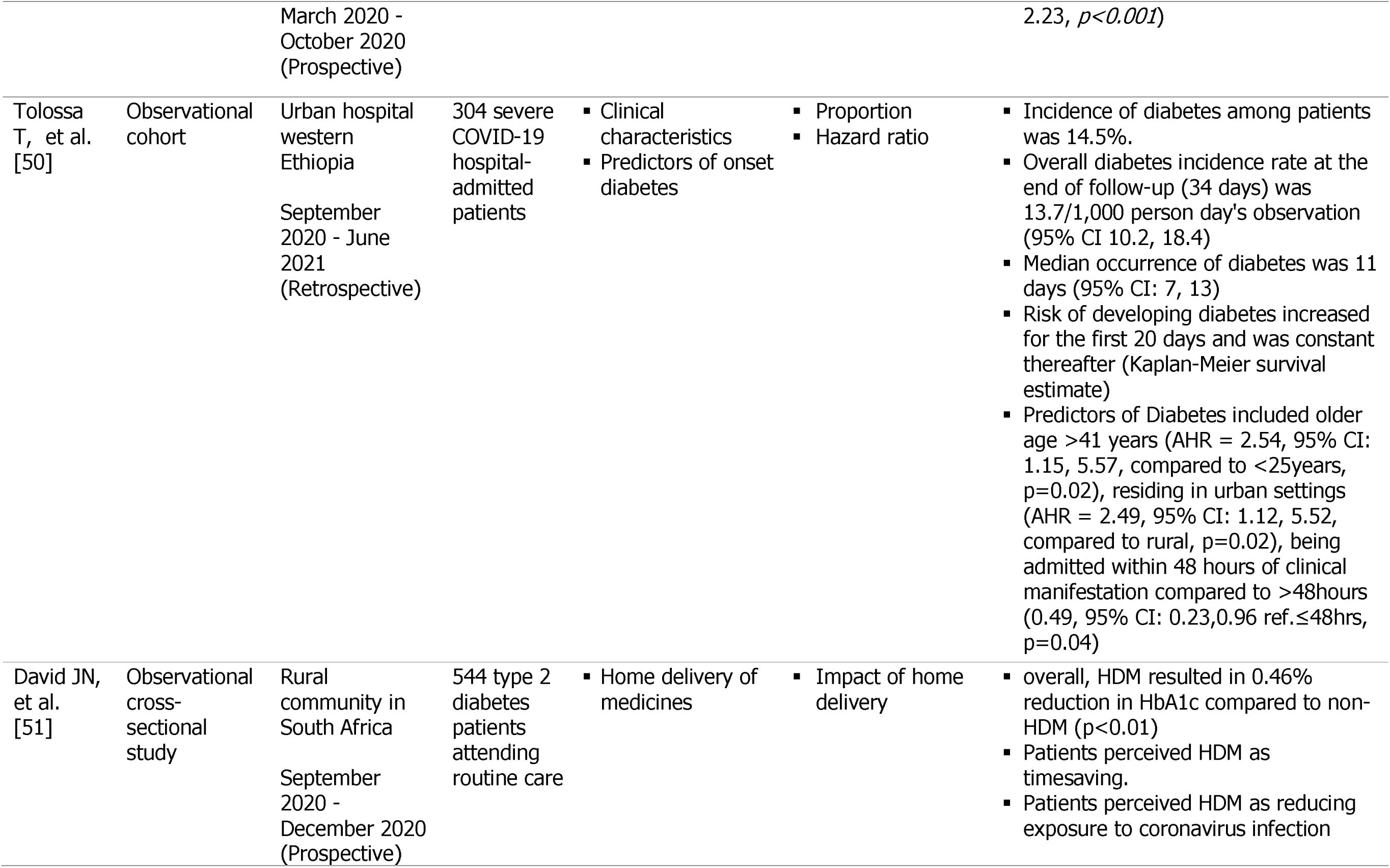
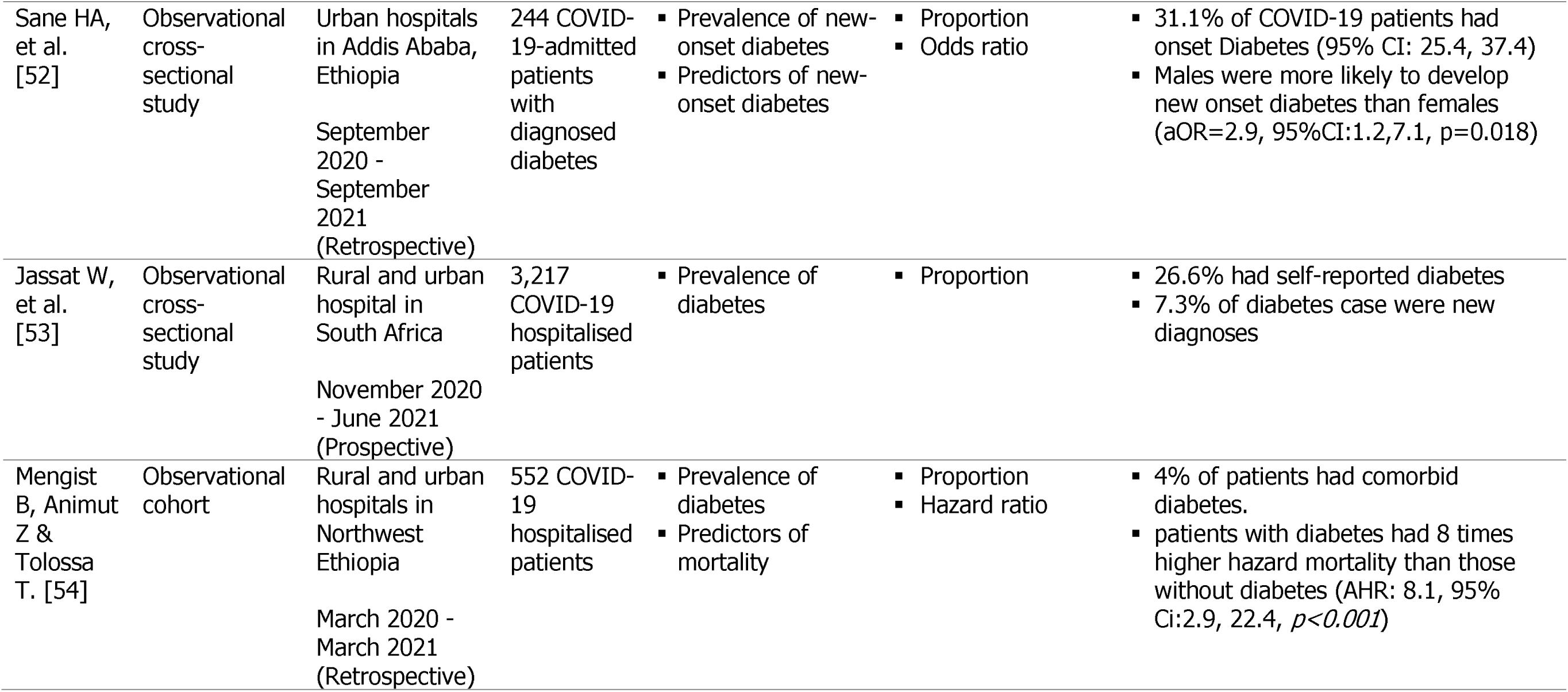
extraction of data from included studies.

The included studies were all observational but dominated by cross-sectional design (69%), with sample sizes ranging from 18 [13] to 3,460,932 [14]. The studies were predominantly retrospective (66.7%) and published between 2021 and 2022 (85.7%). The majority originated in South Africa (40.5%) and were mainly hospital-based (83.3%) and employed quantitative methods (90.4%). The extracted data variables were, DM prevalence among COVID-19 cases, outcomes of DM related to COVID-19 and their predictors, patient-related health management aspects, DM care service delivery aspects, and organisation of DM care related to the pandemic.

### Prevalence and incidence of DM among COVID-19 cases

As shown in Table 2, comorbidity of DM and COVID-19 was very prevalent, with up to 51% pre-existing cases reported, and a mean (SD) figure of 23% (±13.8). Prevalence as high as 31.1% was also reported for new-onset DM among COVID-19 hospitalised cases, and a high incidence rate of 37/1,000 person days [50].

**Table 2:**
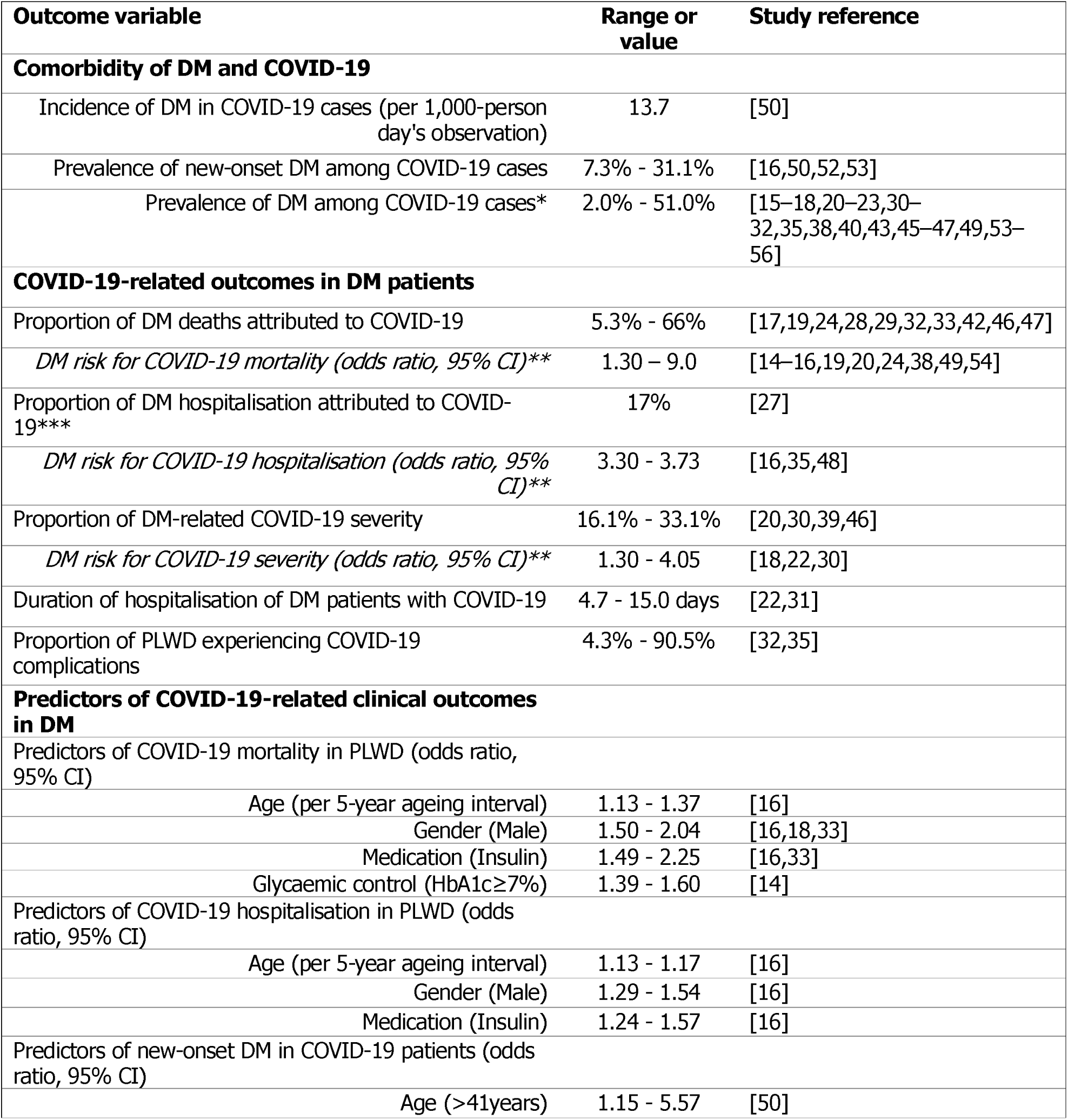
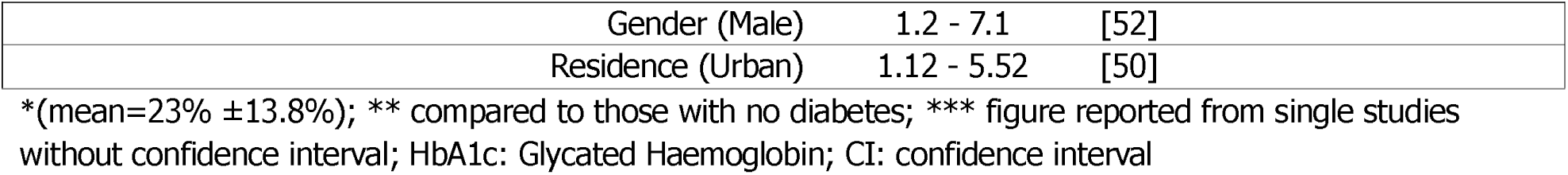
Studies reporting on different COVID-19 outcome variables.

| Outcome variable | Range or value | Study reference |
| --- | --- | --- |
| **Comorbidity of DM and COVID-19** |  |  |
| Incidence of DM in COVID-19 cases (per 1,000-person day's observation) | 13.7 | [50] |
| Prevalence of new-onset DM among COVID-19 cases | 7.3% - 31.1% | [16,50,52,53] |
| Prevalence of DM among COVID-19 cases* | 2.0% - 51.0% | [15–18,20–23,30–32,35,38,40,43,45–47,49,53–56] |
| **COVID-19-related outcomes in DM patients** |  |  |
| Proportion of DM deaths attributed to COVID-19 | 5.3% - 66% | [17,19,24,28,29,32,33,42,46,47] |
| *DM risk for COVID-19 mortality (odds ratio, 95% CI)\*\** | 1.30 – 9.0 | [14–16,19,20,24,38,49,54] |
| Proportion of DM hospitalisation attributed to COVID-19*** | 17% | [27] |
| *DM risk for COVID-19 hospitalisation (odds ratio, 95% CI)\*\** | 3.30 - 3.73 | [16,35,48] |
| Proportion of DM-related COVID-19 severity | 16.1% - 33.1% | [20,30,39,46] |
| *DM risk for COVID-19 severity (odds ratio, 95% CI)\*\** | 1.30 - 4.05 | [18,22,30] |
| Duration of hospitalisation of DM patients with COVID-19 | 4.7 - 15.0 days | [22,31] |
| Proportion of PLWD experiencing COVID-19 complications | 4.3% - 90.5% | [32,35] |
| **Predictors of COVID-19-related clinical outcomes in DM** |  |  |
| Predictors of COVID-19 mortality in PLWD (odds ratio, 95% CI) |  |  |
| Age (per 5-year ageing interval) | 1.13 - 1.37 | [16] |
| Gender (Male) | 1.50 - 2.04 | [16,18,33] |
| Medication (Insulin) | 1.49 - 2.25 | [16,33] |
| Glycaemic control (HbA1c≥7%) | 1.39 - 1.60 | [14] |
| Predictors of COVID-19 hospitalisation in PLWD (odds ratio, 95% CI) |  |  |
| Age (per 5-year ageing interval) | 1.13 - 1.17 | [16] |
| Gender (Male) | 1.29 - 1.54 | [16] |
| Medication (Insulin) | 1.24 - 1.57 | [16] |
| Predictors of new-onset DM in COVID-19 patients (odds ratio, 95% CI) |  |  |
| Age (>41years) | 1.15 - 5.57 | [50] |

|  |  |  |  |
| --- | --- | --- | --- |
|  | Gender (Male) | 1.2 - 7.1 | [52] |
|  | Residence (Urban) | 1.12 - 5.52 | [50] |

\*(mean=23% ±13.8%); \*\* compared to those with no diabetes; \*\*\* figure reported from single studies without confidence interval; HbA1c: Glycated Haemoglobin; CI: confidence interval

### COVID-19-related outcomes of DM and their predictors

As shown in Table 2, mortality, hospitalisation, severity, and complications were the major outcomes related to COVID-19 in DM. The proportions of COVID-19-attributed mortality [17,19,24,28,29,32,33,42,46,47], hospitalisation [27], and severity [20,30,39,46] for PLWD were noticeable to high across the studies. The major predictors of COVID-19-related mortality and hospitalisation in PLWD were age, gender, DM treatment, and glycaemic control. For every 5-year age interval, being male, insulin treatment and HbA1c ≥7.0% were independently associated with higher odds for both COVID-19-related mortality [14,16,18,33] and hospitalisation [16]. On the other hand, new-onset DM, defined as DM diagnosed in hospitalised COVID-19 patients with prior normoglycaemia, was associated with age over 41 years, male gender and urban residence [50,52].

### Impact of the COVID-19 pandemic on DM care

Using an inductive thematic approach, we constructed three major themes from qualitative, mixed methods studies and open-ended quantitative results. The findings were thematically categorised as patient-related health management challenges, DM care service delivery challenges, and reorganisation of DM care delivery (S3 Themes). Table 3 presents a summary of studies that reported on each theme category.

**Table 3:** Studies reporting COVID-19 pandemic’s impact on various aspects of DM care management.

| Major theme | Sub-theme | Reference |
| --- | --- | --- |
| Patient-related health management challenges | Self-management challenges | [15,33,36,37] |
|  | Affordability challenges | [13,37] |
|  | Health service accessibility challenges | [13,37,51] |
| DM care service delivery | Health workforce challenges | [13,34] |
| challenges | Healthcare infrastructure challenges | [13] |
|  | Health information challenges | [25] |
|  | Medicines and medical supplies | [13] |
| Re-organisation of DM care delivery | Patient-level reorganisation of care access | [25,34,51] |
|  | Clinic-level reorganisation of management | [34] |
|  | Community-level re-organisation of community health worker services | [25,34,51] |

### Patient-related health management challenges

The three sub-themes that emerged under patient-related health management challenges were, self-management challenges, affordability challenges, and health service accessibility challenges. Self-management challenges reported among PLWD during the COVID-19 pandemic include reduced daily meal frequency [36,37], inadequate physical activity [26,37], and worsening glycaemic control [15,33,34]. Affordability challenges were related to increased costs of medicines [13] and reduced individual or household income [37]. PLWD also experienced health service accessibility challenges reported as increased clinic waiting time [13] and limited transport means to healthcare facilities [16,51]. Type 1 DM-specific challenges were limited food access, reduced affordability of living costs and accessibility of DM care services [37].

### DM care service delivery challenges

Four sub-themes emerged under DM care delivery challenges, namely, health workforce challenges [13,34], healthcare infrastructure challenges [13,34], health information management challenges [13,25], and medicines and medical supplies [13]. Health workforce challenges were characterised by health workers’ hesitancy towards work and the limited number of available DM specialists. This resulted in fewer active health workers at health facilities that increased workload [13,34]. At the same time, inadequate healthcare infrastructure limited available physical clinic space due to overwhelming patient numbers [13,34]. The COVID-19 pandemic was also characterised by poor management of health information and medical records attributed to the heavy workload of health workers and the fear of the risk of cross-infection while collecting patient data [13,25]. Additionally, the pandemic worsened shortages of medicine and medical supplies at health facilities [13].

### Reorganisation of DM care delivery

Four sub-themes, as shown in Table 3, were categorised under the reorganisation of DM care delivery as a result of the pandemic, namely, patient-level reorganisation of care access [25,34,51], clinic-level reorganisation in management [34], and community-level reorganisation of community health worker services [25,34,51]. The reorganisation of DM care delivery was in response to the challenges patients and healthcare facilities faced in accessing and delivering DM care services. The interventions included delivery of patient medicines to their homes through their community health workers [25,34,51], which addressed the risk of infection and mitigated the health facility accessibility challenges faced by patients during lockdowns [34]. At clinic level, routine non-communicable disease ‘walk-in’ clinics were replaced with a clinic booking system to manage patient appointments and control clinic patient numbers [34]. At the community level, community health workers were empowered to monitor and follow up on patients with non-communicable diseases, including DM, aimed at reducing the workload of health facility staff [25,34,51].

## Discussion

Our scoping review of 42 articles highlighted COVID-19’s impact on DM outcomes and care in SSA. It also lays down existing gaps in knowledge and research. To the best of our knowledge, this is the first systematic scoping review in SSA to investigate outcomes of DM with COVID-19 and the pandemic’s effect on DM care. Our results show an inequitable representation in DM research in countries of SSA, with research outputs mainly contributed by South Africa.

Overall, our scoping review shows that COVID-19 increased the risk of mortality and hospitalisation in PLWD, which were associated with older age, poor glycaemic control, insulin use and being male. These risk factors have also been reported in the US [57], China [58] and the UK [59]. We observed that PLWD had up to nine times higher risk of death, more than three times higher risk of hospitalisation and up to four times higher risk for severity due to COVID-19 compared to those without DM. Notably, similar findings but with varying levels of mortality and morbidity have been reported in China and the USA by Kumar et al. [60]. They revealed higher odds of COVID-19-related mortality (2.16, 95% CI: 1.74-2.65) and severity (2.75, 95% CI: 2.09-3.62) in PLWD than those without DM. COVID-19’s impact on DM clinical outcomes in SSA is significant and consistent with reports from the World Health Organization that indicate COVID-19 is deadlier in PLWD in Africa due to the region’s characteristic poor glycaemic control [61,62]. Additionally, COVID-19 was associated with an increased risk of developing new-onset DM, especially among hospitalised COVID cases over 41 years, males and urban residents. We observed a DM incident rate of 13.7/1,000 person-days (the equivalent of 5/1,000 person-years) and a prevalence of new-onset DM of up to 31% among COVID-19 cases in SSA. This rate is, however, considerably lower than what has been reported in the US (23-83/1,000 person-years) [63], England (37.2/1,000 person-years) [64] and China (13.5/1,000 person-years) [65]. Whereas the variation in diabetes incidence among COVID-19 patients in SSA may be due to underreporting, COVID-19’s epidemiological threat to the growing burden of DM in SSA needs to be tracked.

As a pandemic, COVID-19 also impacted DM indirectly by causing disruptions in patient self-management routines and delivery of DM services in SSA. As our scoping review highlights, this impact manifested through challenges posed by instituted COVID-19 restrictions. For PLWD, we observe that this negatively affected their dietary intake and engagement in physical activity and limited their access to healthcare. The experience in SSA was however, in marked contrast with reports from India [66] and the UK [67], which showed no notable negative COVID-19 impact on access to essential services among PLWD. This stark variation may be explained by the different countries’ approaches to containing COVID-19, which in most SSA countries mainly targeted geographical containment, closure of non-essential services and prohibition of gatherings [68]. These unprecedented approaches created blockades to accessibility and affordability of various services, including health and social services [69–71]. On other grounds, there was a considerable shortage of health workforce, physical infrastructure and severe shortages of DM medicine and medical supplies. Whereas we acknowledge the pre-existence of challenges in the health workforce, healthcare infrastructure and medical supplies in SSA before the COVID-19 pandemic, the magnitude might have worsened during the pandemic due to a shift in healthcare resource prioritisation toward COVID-19 [72][73][74][73,75].

Interestingly, we also observed from our scoping review that the pandemic presented some opportunities for DM care innovation. For instance, the delivery of medicine to patient homes implemented in South Africa reportedly reduced the risk of COVID-19 infection among PLWD, mitigated DM care access challenges and ensured continued chronic patient follow-up [40]. Home delivery of medicine has also been reported to improve treatment adherence among chronic disease patients in Rwanda, which shows its feasibility in other SSA countries [76]. The pandemic, as demonstrated in South Africa, has also evidenced the value of integrating chronic non-communicable disease prevention and care in the services of community health workers. Additionally, clinic booking systems introduced to replace walk-in clinics in public health centres were found to mitigate clinic overcrowding, reduce clinic waiting time, and provide better doctor-to-patient time. These changes in the reorganisation of healthcare service delivery proved vital in addressing many challenges posed by the COVID-19 pandemic and offer lessons to policy and practice in future planning.

### Gaps in knowledge and research

In our scoping review, we note various gaps in knowledge that can inform subsequent research. Firstly, there is a gap in the published literature on the use of guidelines for managing COVID-19 and DM in SSA countries, which would help evaluate their appropriateness for future similar occurrences. Secondly, the studies in our scoping review did not report on vaccine uptake or how the different ‘waves’ of COVID-19 infection influenced COVID-19 outcomes among PLWD. This would provide an understanding of the outcomes of PLWD across evolving pandemic dynamics and health system interventions. Exploiting research opportunities to address such gaps in knowledge can provide further and comprehensive understanding to shape appropriate post-pandemic DM care approaches and health system preparedness in addressing chronic care vulnerabilities during possible future pandemics.

### Limitations

While this scoping review provides reliable information by scoping various research types and sources, there are some limitations. Firstly, our scoping review only included articles published in English. This may have limited studies published from non-English speaking countries within SSA; therefore, some relevant studies may have been missed. However, considering what was retrieved from most SSA countries, we predict this number to be likely minimal. Secondly, the included studies were dominated by three countries, which may limit the generalisation of findings to SSA. Thirdly, the studies were mainly conducted in the initial phase of the pandemic in 2020, indicating that changes experienced after that may render some findings unrepresentative of the post-2020 dynamics including the impact of emerging COVID-19 variants. Moreover, the limited disaggregation of data by studies in our scoping review, especially age, gender and type of DM, limited the drawing of specific conclusions and analyses. Finally, we only included peer-reviewed literature, which may have excluded some valuable literature sources such as manuscripts, institutional reports and archives. Nevertheless, this scoping review provided critical information and insights on how COVID-19 impacted PLWD and healthcare systems in SSA.

## Conclusions

COVID-19 increased mortality and morbidity among PLWD and the occurrence of DM. In addition, the pandemic worsened DM self-care and DM service delivery generally. Therefore, further research in SSA is needed to understand the disease syndemism of pandemics such as COVID-19 and DM to inform future management strategies and policy considerations.

## Supporting information

Search Strategy

Themes

## Data Availability

All data produced in the present work are contained in the manuscript

## Supporting information

S1 PRISMA-ScR Checklist

S2 Search strategy

S3 Themes

## References

1. Wang H, Paulson KR, Pease SA, Watson S, Comfort H, Zheng P, et al. Estimating excess mortality due to the COVID-19 pandemic: a systematic analysis of COVID-19-related mortality, 2020–21. Lancet. 2022;399: 1513–1536. doi:10.1016/S0140-6736(21)02796-3

2. Post LA, Argaw ST, Jones C, Moss CB, Resnick D, Singh LN, et al. A SARS-CoV-2 Surveillance System in Sub-Saharan Africa: Modeling Study for Persistence and Transmission to Inform Policy. J Med Internet Res. 2020;22: e24248. doi:10.2196/24248

3. Fu Y, Hu L, Ren H-W, Zuo Y, Chen S, Zhang Q-S, et al. Prognostic Factors for COVID-19 Hospitalized Patients with Preexisting Type 2 Diabetes. Merlotti D, editor. Int J Endocrinol. 2022;2022: 1–13. doi:10.1155/2022/9322332

4. Hayden MR. An Immediate and Long-Term Complication of COVID-19 May Be Type 2 Diabetes Mellitus: The Central Role of β-Cell Dysfunction, Apoptosis and Exploration of Possible Mechanisms. Cells. 2020;9: 2475. doi:10.3390/cells9112475

5. Landstra CP, de Koning EJP. COVID-19 and Diabetes: Understanding the Interrelationship and Risks for a Severe Course. Front Endocrinol (Lausanne). 2021;12. doi:10.3389/fendo.2021.649525

6. Filip R, Gheorghita Puscaselu R, Anchidin-Norocel L, Dimian M, Savage WK. Global Challenges to Public Health Care Systems during the COVID-19 Pandemic: A Review of Pandemic Measures and Problems. J Pers Med. 2022;12: 1295. doi:10.3390/jpm12081295

7. Topkar V. Interactions Between Diabetes And Covid-19: A Scoping Review. Yale University. 2022. Available: https://elischolar.library.yale.edu/ysphtdl/2207?utm_source=elischolar.library.yale.edu%2Fysphtdl%2F2207&utm_medium=PDF&utm_campaign=PDFCoverPages

8. Khunti K, Aroda VR, Aschner P, Chan JCN, Del Prato S, Hambling CE, et al. The impact of the COVID-19 pandemic on diabetes services: planning for a global recovery. Lancet Diabetes Endocrinol. 2022;10: 890–900. doi:10.1016/S2213-8587(22)00278-9

9. IDF. IDF Diabetes Atlas 10th Edition. Brussels; 2021.

10. Cooke A, Smith D, Booth A. Beyond PICO. Qual Health Res. 2012;22: 1435–1443. doi:10.1177/1049732312452938

11. Richardson S, Wilson MC, Nishikawa J, Hayward RS. The well-built clinical question: a key to evidence-based decisions. ACP J Club. 1995;123.

12. World Bank. FOCUS: Sub-Saharan Africa. 2021 [cited 22 Feb 2024]. Available: https://openknowledge.worldbank.org/pages/focus-sub-saharan-africa

13. Tagoe ET, Nonvignon J, van Der Meer R, Megiddo I, Godman B. Challenges to the delivery of clinical diabetes services in Ghana created by the COVID-19 pandemic. J Health Serv Res Policy. 2023;28: 58–65. doi:10.1177/13558196221111708

14. Boulle A, Davies M-A, Hussey H, Ismail M, Morden E, Vundle Z, et al. Risk Factors for Coronavirus Disease 2019 (COVID-19) Death in a Population Cohort Study from the Western Cape Province, South Africa. Clin Infect Dis. 2021;73: e2005–e2015. doi:10.1093/cid/ciaa1198

15. Mash RJ, Presence-Vollenhoven M, Adeniji A, Christoffels R, Doubell K, Eksteen L, et al. Evaluation of patient characteristics, management and outcomes for COVID-19 at district hospitals in the Western Cape, South Africa: descriptive observational study. BMJ Open. 2021;11: e047016. doi:10.1136/bmjopen-2020-047016

16. Dave JA, Tamuhla T, Tiffin N, Levitt NS, Ross IL, Toet W, et al. Risk factors for COVID-19 hospitalisation and death in people living with diabetes: A virtual cohort study from the Western Cape Province, South Africa. Diabetes Res Clin Pract. 2021;177: 108925. doi:10.1016/j.diabres.2021.108925

17. van Hoving DJ, Hattingh N, Pillay SK, Lockey T, McAlpine DJ, Nieuwenhuys K, et al. Demographics and clinical characteristics of hospitalised patients under investigation for COVID-19 with an initial negative SARS-CoV-2 PCR test result. African J Emerg Med. 2021;11: 429–435. doi:10.1016/j.afjem.2021.09.002

18. Ratshikhopha E, Muvhali M, Naicker N, Tlotleng N, Jassat W, Singh T. Disease Severity and Comorbidities among Healthcare Worker COVID-19 Admissions in South Africa: A Retrospective Analysis. Int J Environ Res Public Health. 2022;19: 5519. doi:10.3390/ijerph19095519

19. Claassen N, van Wyk G, van Staden S, Basson MMD. Experiencing COVID-19 at a large district level hospital in Cape Town: A retrospective analysis of the first wave. South African J Infect Dis. 2022;37. doi:10.4102/sajid.v37i1.317

20. Abraha HE, Gessesse Z, Gebrecherkos T, Kebede Y, Weldegiargis AW, Tequare MH, et al. Clinical features and risk factors associated with morbidity and mortality among patients with COVID-19 in northern Ethiopia. Int J Infect Dis. 2021;105: 776–783. doi:10.1016/j.ijid.2021.03.037

21. Kaswa R, Yogeswaran P, Cawe B. Clinical outcomes of hospitalised COVID-19 patients at Mthatha Regional Hospital, Eastern Cape, South Africa: A retrospective study. South African Fam Pract. 2021;63. doi:10.4102/safp.v63i1.5253

22. Fouda Mbarga N, Epee E, Mbarga M, Ouamba P, Nanda H, Nkengni A, et al. Clinical profile and factors associated with COVID-19 in Yaounde, Cameroon: A prospective cohort study. Zivkovic AR, editor. PLoS One. 2021;16: e0251504. doi:10.1371/journal.pone.0251504

23. Kwaghe VG, Habib ZG, Akor AA, Thairu Y, Bawa A, Adebayo FO, et al. Clinical characteristics and outcome of the first 200 patients hospitalized with coronavirus disease-2019 at a treatment center in Abuja, Nigeria: a retrospective study. Pan Afr Med J. 2022;41: 118. doi:10.11604/pamj.2022.41.118.26594

24. Leulseged TW, Maru EH, Hassen IS, Zewde WC, Chamiso NH, Abebe DS, et al. Predictors of death in severe COVID-19 patients at millennium COVID-19 care center in Ethiopia: a case-control study. Pan Afr Med J. 2021;38. doi:10.11604/pamj.2021.38.351.28831

25. Brey Z, Mash R, Goliath C, Roman D. Home delivery of medication during Coronavirus disease 2019, Cape Town, South Africa: Short report. African J Prim Heal Care Fam Med. 2020;12. doi:10.4102/phcfm.v12i1.2449

26. Abate HK, Ferede YM, Mekonnen CK. Adherence to physical exercise recommendations among type 2 diabetes patients during the COVID-19 pandemic. Int J Africa Nurs Sci. 2022;16: 100407. doi:10.1016/j.ijans.2022.100407

27. Bepouka BI, Mandina M, Makulo JR, Longokolo M, Odio O, Mayasi N, et al. Predictors of mortality in COVID-19 patients at Kinshasa University Hospital, Democratic Republic of the Congo (from March to June 2020). Pan Afr Med J. 2020;37. doi:10.11604/pamj.2020.37.105.25279

28. Poaty H, Emergence Poaty G, NDziessi G, Godefroy Ngakeni E, Doukaga Makouka T, Soussa Gadoua R, et al. Diabetes and COVID-19 in Congolese patients. Afr Health Sci. 2021;21: 1100–1106. doi:10.4314/ahs.v21i3.18

29. Ikram A, Pillay S. Hyperglycaemia, diabetes mellitus and COVID-19 in a tertiary hospital in KwaZulu-Natal. J Endocrinol Metab Diabetes South Africa. 2022;27: 32–41. doi:10.1080/16089677.2021.1997427

30. Leulseged TW, Abebe KG, Hassen IS, Maru EH, Zewde WC, Chamiso NW, et al. COVID-19 disease severity and associated factors among Ethiopian patients: A study of the millennium COVID-19 care center. Taghizadeh-Hesary F, editor. PLoS One. 2022;17: e0262896. doi:10.1371/journal.pone.0262896

31. Leulseged TW, Hassen IS, Maru EH, Zewsde WC, Chamiso NW, Bayisa AB, et al. Characteristics and outcome profile of hospitalized African patients with COVID-19: The Ethiopian context. Mossong J, editor. PLoS One. 2021;16: e0259454. doi:10.1371/journal.pone.0259454

32. Adjei P, Afriyie-Mensah J, J. Ganu V, Puplampu P, Opoku-Asare B, Dzefi-Tettey K, et al. Clinical characteristics of COVID-19 patients admitted at the Korle-Bu Teaching Hospital, Accra, Ghana. Ghana Med J. 2020;54: 33–38. doi:10.4314/gmj.v54i4s.6

33. Van der Westhuizen J-N, Hussey N, Zietsman M, Salduker N, Manning K, Dave JA, et al. Low mortality of people living with diabetes mellitus diagnosed with COVID-19 and managed at a field hospital in Western Cape Province, South Africa. South African Med J. 2021;111: 961. doi:10.7196/SAMJ.2021.v111i10.15779

34. Delobelle PA, Abbas M, Datay I, De Sa A, Levitt N, Schouw D, et al. Non-communicable disease care and management in two sites of the Cape Town Metro during the first wave of COVID-19: A rapid appraisal. African J Prim Heal Care Fam Med. 2022;14. doi:10.4102/phcfm.v14i1.3215

35. Crankson S, Pokhrel S, Anokye NK. Determinants of COVID-19-Related Length of Hospital Stays and Long COVID in Ghana: A Cross-Sectional Analysis. Int J Environ Res Public Health. 2022;19: 527. doi:10.3390/ijerph19010527

36. Ephraim RKD, Duah E, Nkansah C, Amoah S, Fosu E, Afrifa J, et al. Psychological impact of COVID-19 on diabetes mellitus patients in Cape Coast, Ghana: a cross-sectional study. Pan Afr Med J. 2021;40: 76. doi:10.11604/pamj.2021.40.76.26834

37. Habineza JC, James S, Sibomana L, Klatman E, Uwingabire E, Maniam J, et al. Perceived impact of the COVID-19 pandemic on young adults with type 1 diabetes in Rwanda. Pan Afr Med J. 2021;40. doi:10.11604/pamj.2021.40.252.28899

38. Baguma S, Okot C, Alema NO, Apiyo P, Layet P, Acullu D, et al. Factors Associated With Mortality Among the COVID-19 Patients Treated at Gulu Regional Referral Hospital: A Retrospective Study. Front Public Heal. 2022;10. doi:10.3389/fpubh.2022.841906

39. Iroungou BA, Mangouka LG, Bivigou-Mboumba B, Moussavou-Boundzanga P, Obame-Nkoghe J, Nzigou Boucka F, et al. Demographic and Clinical Characteristics Associated With Severity, Clinical Outcomes, and Mortality of COVID-19 Infection in Gabon. JAMA Netw Open. 2021;4: e2124190. doi:10.1001/jamanetworkopen.2021.24190

40. Emmanuel Awucha N, Chinelo Janefrances O, Chima Meshach A, Chiamaka Henrietta J, Ibilolia Daniel A, Esther Chidiebere N. Impact of the COVID-19 Pandemic on Consumers’ Access to Essential Medicines in Nigeria. Am J Trop Med Hyg. 2020;103: 1630–1634. doi:10.4269/ajtmh.20-0838

41. Kaswa RP, B Meel. A Study on the Characteristic Features of Covid-19 Deaths in a Regional Hospital in Mthatha in the Eastern Cape, South Africa. Indian J Forensic Med Toxicol. 2021;16: 1554–1559. doi:10.37506/ijfmt.v16i1.17723

42. Usui R, Kanamori S, Aomori M, Watabe S. Analysis of COVID-19 mortality in patients with comorbidities in Côte d’Ivoire. J Public Health Africa. 2022;13. doi:10.4081/jphia.2022.1748

43. Sikhosana ML, Jassat W, Makatini Z. Characteristics of hospitalised COVID-19 patients during the first two pandemic waves, Gauteng. South African J Infect Dis. 2022;37. doi:10.4102/sajid.v37i1.434

44. Elijah IM, Amsalu E, Jian X, Cao M, Mibei EK, Kerosi DO, et al. Characterization and determinant factors of critical illness and in-hospital mortality of COVID-19 patients: A retrospective cohort of 1,792 patients in Kenya. Biosaf Heal. 2022;4: 330–338. doi:10.1016/j.bsheal.2022.06.002

45. Hardy YO, Libhaber E, Ofori E, Amenuke DAY, Kontoh SA, Dankwah JA, et al. Clinical and laboratory profile and outcomes of hospitalized COVID□19 patients with type 2 diabetes mellitus in Ghana – A single□center study. Endocrinol Diabetes Metab. 2023;6. doi:10.1002/edm2.391

46. Huluka DK, Etissa EK, Ahmed S, Abule HA, Getachew N, Abera S, et al. Clinical Characteristics and Treatment Outcomes of COVID-19 Patients at Eka Kotebe General Hospital, Addis Ababa, Ethiopia. Am J Trop Med Hyg. 2022;107: 252–259. doi:10.4269/ajtmh.21-1270

47. Nyasulu PS, Ayele BT, Koegelenberg CF, Irusen E, Lalla U, Davids R, et al. Clinical characteristics associated with mortality of COVID-19 patients admitted to an intensive care unit of a tertiary hospital in South Africa. Aouissi HA, editor. PLoS One. 2022;17: e0279565. doi:10.1371/journal.pone.0279565

48. Solanki G, Wilkinson T, Bansal S, Shiba J, Manda S, Doherty T. COVID-19 hospitalization and mortality and hospitalization-related utilization and expenditure: Analysis of a South African private health insured population. Kuo RN, editor. PLoS One. 2022;17: e0268025. doi:10.1371/journal.pone.0268025

49. Diarra M, Barry A, Dia N, Diop M, Sonko I, Sagne S, et al. First wave COVID-19 pandemic in Senegal: Epidemiological and clinical characteristics. Mossong J, editor. PLoS One. 2022;17: e0274783. doi:10.1371/journal.pone.0274783

50. Tolossa T, Lema M, Wakuma B, Turi E, Fekadu G, Mulisa D, et al. Incidence and predictors of diabetes mellitus among severe COVID-19 patients in western Ethiopia: a retrospective cohort study. J Endocrinol Metab Diabetes South Africa. 2023;28: 42–48. doi:10.1080/16089677.2022.2144016

51. David NJ, Bresick G, Moodaley N, Von Pressentin KB. Measuring the impact of community-based interventions on type 2 diabetes control during the COVID-19 pandemic in Cape Town – A mixed methods study. South African Fam Pract. 2022;64. doi:10.4102/safp.v64i1.5558

52. Sane AH, Mekonnen MS, Tsegaw MG, Zewde WC, Mesfin EG, Beyene HA, et al. New Onset of Diabetes Mellitus and Associated Factors among COVID-19 Patients in COVID-19 Care Centers, Addis Ababa, Ethiopia 2022. Kretchy I, editor. J Diabetes Res. 2022;2022: 1–9. doi:10.1155/2022/9652940

53. Jassat W, Mudara C, Vika C, Dryden M, Masha M, Arendse T, et al. Undiagnosed comorbidities among individuals hospitalised with COVID-19 in South African public hospitals. South African Med J. 2022;112: 747–752. doi:10.7196/SAMJ.2022.v112i9.16417

54. Mengist B, Animut Z, Tolossa T. Incidence and predictors of mortality among COVID-19 patients admitted to treatment centers in North West Ethiopia; A retrospective cohort study, 2021. Int J Africa Nurs Sci. 2022;16: 100419. doi:10.1016/j.ijans.2022.100419

55. Kaswa P, Meel B. A Study on the Characteristic Features of Covid-19 Deaths in a Regional Hospital in Mthatha in the Eastern Cape, South Africa. Indian J Forensic Med Toxicol. 2022;16. doi:10.37506/ijfmt.v16i1.17723

56. Elijah IM, Amsalu E, Jian X, Cao M, Mibei EK, Kerosi DO, et al. Characterization and determinant factors of critical illness and in-hospital mortality of COVID-19 patients: A retrospective cohort of 1,792 patients in Kenya. Biosaf Heal. 2022;4: 330–338. 10.1016/j.bsheal.2022.06.002

57. Chen U-I, Xu H, Krause TM, Greenberg R, Dong X, Jiang X. Factors Associated With COVID-19 Death in the United States: Cohort Study. JMIR Public Heal Surveill. 2022;8: e29343. doi:10.2196/29343

58. Xu PP, Tian RH, Luo S, Zu ZY, Fan B, Wang XM, et al. Risk factors for adverse clinical outcomes with COVID-19 in China: a multicenter, retrospective, observational study. Theranostics. 2020;10: 6372–6383. doi:10.7150/thno.46833

59. Bhaskaran K, Bacon S, Evans SJ, Bates CJ, Rentsch CT, MacKenna B, et al. Factors associated with deaths due to COVID-19 versus other causes: population-based cohort analysis of UK primary care data and linked national death registrations within the OpenSAFELY platform. Lancet Reg Heal -Eur. 2021;6: 100109. doi:10.1016/j.lanepe.2021.100109

60. Kumar A, Arora A, Sharma P, Anikhindi SA, Bansal N, Singla V, et al. Is diabetes mellitus associated with mortality and severity of COVID-19? A meta-analysis. Diabetes Metab Syndr Clin Res Rev. 2020;14: 535–545. doi:10.1016/j.dsx.2020.04.044

61. Burki T. COVID-19 and diabetes in Africa: a lethal combination. Lancet Diabetes Endocrinol. 2022;10: 23. doi:10.1016/S2213-8587(21)00315-6

62. Fina Lubaki J-P, Omole OB, Francis JM. Glycaemic control among type 2 diabetes patients in sub-Saharan Africa from 2012 to 2022: a systematic review and meta-analysis. Diabetol Metab Syndr. 2022;14: 134. doi:10.1186/s13098-022-00902-0

63. Birabaharan M, Kaelber DC, Pettus JH, Smith DM. Risk of New-Onset Type 2 Diabetes Mellitus in 600,055 Persons after COVID-19: a cohort study. Diabetes, Obes Metab. 2022;24: 1176–1179. doi:10.1111/dom.14659

64. Tazare J, Walker AJ, Tomlinson LA, Hickman G, Rentsch CT, Williamson EJ, et al. Rates of serious clinical outcomes in survivors of hospitalisation with COVID-19 in England: a descriptive cohort study within the OpenSAFELY platform. Wellcome Open Res. 2022;7: 142. doi:10.12688/wellcomeopenres.17735.1

65. Xie Y, Al-Aly Z. Risks and burdens of incident diabetes in long COVID: a cohort study. lancet Diabetes Endocrinol. 2022;10: 311–321. doi:10.1016/S2213-8587(22)00044-4

66. Madan J, Blonquist T, Rao E, Marwaha A, Mehra J, Bharti R, et al. Effect of COVID-19 Pandemic-Induced Dietary and Lifestyle Changes and Their Associations with Perceived Health Status and Self-Reported Body Weight Changes in India: A Cross-Sectional Survey. Nutrients. 2021;13: 3682. doi:10.3390/nu13113682

67. O’Connell M, Smith K, Stroud R. The dietary impact of the COVID-19 pandemic. J Health Econ. 2022;84: 102641. doi:10.1016/j.jhealeco.2022.102641

68. Haider N, Osman AY, Gadzekpo A, Akipede GO, Asogun D, Ansumana R, et al. Lockdown measures in response to COVID-19 in nine sub-Saharan African countries. BMJ Glob Heal. 2020;5: e003319. doi:10.1136/bmjgh-2020-003319

69. Sseguya W, James S, Manfred B, Munyagwa M, Klatman E, Ogle G, et al. Impact of COVID-19 pandemic on young persons with type 1 diabetes in western Uganda. Manuscr Submitt Publ. 2021.

70. Kebirungi H, Mwenyango H. Impacts of COVID-19 Pandemic Lockdown on the Livelihoods of Male Commercial Boda-Boda Motorists in Uganda. In: Laituri M, Richardson RB, Kim J, editors. The Geographies of COVID-19: Geospatial Stories of a Global Pandemic. Cham: Springer International Publishing; 2022. pp. 195–207. doi:10.1007/978-3-031-11775-6_16

71. Hrynick TA, Ripoll Lorenzo S, Carter SE. COVID-19 response: mitigating negative impacts on other areas of health. BMJ Glob Heal. 2021;6: e004110. doi:10.1136/bmjgh-2020-004110

72. Uwizeyimana T, Hashim HT, Kabakambira JD, Mujyarugamba JC, Dushime J, Ntacyabukura B, et al. Drug supply situation in Rwanda during COVID-19: issues, efforts and challenges. J Pharm Policy Pract. 2021;14: 12. doi:10.1186/s40545-021-00301-2

73. Amu H, Dowou RK, Saah FI, Efunwole JA, Bain LE, Tarkang EE. COVID-19 and Health Systems Functioning in Sub-Saharan Africa Using the “WHO Building Blocks”: The Challenges and Responses. Front Public Heal. 2022;10. doi:10.3389/fpubh.2022.856397

74. Moolla I, Hiilamo H. Health system characteristics and COVID-19 performance in high-income countries. BMC Health Serv Res. 2023;23: 244. doi:10.1186/s12913-023-09206-z

75. Ayanore MA, Amuna N, Aviisah M, Awolu A, Kipo-Sunyehzi DD, Mogre V, et al. Towards Resilient Health Systems in Sub-Saharan Africa: A Systematic Review of the English Language Literature on Health Workforce, Surveillance, and Health Governance Issues for Health Systems Strengthening. Ann Glob Heal. 2019;85. doi:10.5334/aogh.2514

76. Tran DN, Kangogo K, Amisi JA, Kamadi J, Karwa R, Kiragu B, et al. Community-based medication delivery program for antihypertensive medications improves adherence and reduces blood pressure. Weinrauch LA, editor. PLoS One. 2022;17: e0273655. doi:10.1371/journal.pone.0273655

