## Supplementary material for "Impact of COVID-19 on diabetes mellitus outcomes and care in sub-Saharan Africa: A scoping review": Search Strategy

**S2: DETAILED SEARCH STRATEGY**

**Summary of keywords developed using the SPIDER and PICO elements to guide database searches.**

|  | **Keywords of interest** |
| --- | --- |
| **Sample/*Population*** | Persons with diabetes OR persons with COVID-19 OR persons with SARS-CoV-2 OR health workers OR community health workers OR policymakers |
| **Phenomenon of Interest / *Intervention*** | Coronavirus OR SARS-CoV-2 OR COVID-19 OR COVID-19 pandemic OR diabetes mellitus OR Diabetes OR diabetes care |
| **Design** | survey OR questionnaire OR interview OR focus group discussion OR observation OR randomized control trial OR cases study |
| **Evaluation /Outcome** | treatment outcome OR intervention outcome OR predictors of outcome OR experiences OR opinions OR views OR attitudes OR perceptions |
| **Research type** | qualitative OR mixed methods OR quantitative |

**Database-specific search strategy**

1. **Medline**

| Database access link | Via PubMed: <https://pubmed.ncbi.nlm.nih.gov/?term=> |
| --- | --- |
| Search terms | Coronavirus; COVID-19  Diabetes mellitus  Africa south of the Sahara  Africa |
| Search String | ((("coronavirus"[MeSH Terms]) OR ("covid 19"[MeSH Terms])) AND ("diabetes mellitus") AND (africa[MeSH Terms]) |
| Search limits/Filters | Year of publication: 1 Jan 2020 - Date |
| Date of search | Initial: 08 May 2022  Updated: 21 March 2023 |
| Number of articles retrieved | 96 |
| Search notes | An initial search with "Africa south of the Sahara [MeSH]" yielded 48 results, 23 less than the search with Africa. On checking both results, some articles from "Africa" covered countries from sub-Saharan Africa, therefore finding it more appropriate |

2. **Web of Science**

| Database access link | <https://www.webofscience.com/wos/woscc/advanced-search> |
| --- | --- |
| Search terms | Coronavirus  COVID-19  SARS-CoV-2  Diabetes mellitus  Africa |
| Search String | ((((ALL=(COVID-19)) OR ALL=(coronavirus)) OR ALL=(SARS-Cov 2)) AND ALL=(Diabetes mellitus)) AND ALL=(Africa)  Query link: <https://www.webofscience.com/wos/woscc/summary/c0b19350-f627-492a-9af8-2c83a2470d0a-36805e01/relevance/1> |
| Search limits/Filters | Open |
| Date of search | Initial: 08/05/2022  Updated: 21/03/2023 |
| Number of articles retrieved | 79 |
| Search notes | NA |

3. **Cumulative Index to Nursing and Allied Health Libraries** (CINAHL)

| Database access link | Via EBSCO: <https://web.p.ebscohost.com/ehost/search/advanced?vid=0&sid=61f0233a-abd5-41f6-b664-213f6601c510%40redis> |
| --- | --- |
| Search terms | COVID-19  Diabetes mellitus  Africa |
| Search String | TX covid-19 AND TX diabetes mellitus AND TX africa |
| Search limits/Filters | Year of publication: 1 Jan 2020 to Date |
| Date of search | Initial: 09/05/2022  Updated: 21/03/2023 |
| Number of articles retrieved | 22 |
| Search notes | Advanced search using "All Text" (TX) |

4. **African Index Medicus** (AIM)

| Database access link | <https://pesquisa.bvsalud.org/gim/decs-locator/?lang=en> |
| --- | --- |
| Search terms | Coronavirus  COVID-19  SARS-CoV-2  Diabetes mellitus  Diabetes, Africa |
| Search String | (tw:(coronavirus)) AND (tw:(diabetes)) |
| Search limits/Filters | Filter:  Database: AIM  Language: English |
| Date of search | Initial: 09/05/2022  Updated: 21/03/2023 |
| Number of articles retrieved | 22 |
| Search notes | African Index Medicus (AIM) was accessed via GIM that was advance searched using "title, abstract and subject" fields i.e. tw  search details: tw:((tw:(coronavirus)) AND (tw:(diabetes))) AND (collection_gim:("AIM") AND la:("en")) |

5. **Google Scholar**

| Database access link | <https://scholar.google.com/#d=gs_asd> |
| --- | --- |
| Search terms | Coronavirus  COVID-19  SARS-CoV-2  Diabetes  Diabetes mellitus |
| Search String | coronavirus and diabetes |
| Search limits/Filters | with the exact phrase;  anywhere in the article; dates (2020 - 2023) |
| Date of search | Initial: 09/05/2022  Updated: 21/03/2023 |
| Number of articles retrieved | 89 |
| Search notes | Performed initial multiple searches using different search term combinations and going through each set of output to see if returned results met the search objective. The database has a simple interface with limited flexibility of search and filter options. Filtering was manually done (for Africa region and articles) |

6. **Cochrane Library**

| Database access link | <https://covid-19.cochrane.org/> |
| --- | --- |
| Search terms | Coronavirus  COVID-19  SARS-CoV-2  Diabetes mellitus  Diabetes; Africa |
| Search String | "Africa" and "Diabetes" |
| Search limits/Filters | Filtered for "Africa" and " Diabetes" in the Cochrane COVID-19 Study register of the Cochrane Library |
| Date of search | Initial: 09/05/2022  Updated: 22/03/2023 |
| Number of articles retrieved | 126 |
| Search notes | Via the Cochrane Library, accessed was gained to the Cochrane COVID-19 study register which is the collection of all trials and studies related to COVID-19.  Link: https://covid-19.cochrane.org/ |

7. **Scopus**

| Database access link | Via Elsevier: <https://www.scopus.com/search/form.uri?display=basic#basic> |
| --- | --- |
| Search terms | Coronavirus  COVID-19  SARS-CoV-2  Diabetes  Diabetes mellitus  Countries south of the Sahara |
| Search String | coronavirus OR COVID AND Diabetes |
| Search limits/Filters | Countries in sub-Saharan Africa; Published 2020 - Present |
| Date of search | Initial: 10/05/2022  Updated: 22/03/2023 |
| Number of articles retrieved | 18 |
| Search notes | NA |

8. **Education Resource Information Centre (ERIC)**

| Database access link | Via EBSCO: <https://web.p.ebscohost.com/ehost/search/advanced?vid=0&sid=1d74b95f-b206-46f6-bad2-c1c321288611%40redis> |
| --- | --- |
| Search terms | Coronavirus  COVID-19  SARS-CoV-2  Diabetes mellitus  Diabetes  Africa |
| Search String | AB coronavirus OR AB covid-19 AND AB diabetes mellitus [searched Abstract] |
| Search limits/Filters | Apply related words; Apply equivalent subjects; Full-text availability; sub-Saharan Africa; published 01 Jan 2020 - 2023 |
| Date of search | Initial: 10/05/2022  Updated: 22/03/2023 |
| Number of articles retrieved | 05 |
| Search notes | Searched using Boolean operators; Apply related words; Apply equivalent subjects; Full-text availability; sub-Saharan Africa; published 01 Jan 2020 - 2023 |

9. **Science Direct**

| Database access link | Via Elsevier: <https://www.sciencedirect.com/search> |
| --- | --- |
| Search terms | Coronavirus  COVID-19  SARS-CoV-2  Diabetes  Diabetes Mellitus  Africa  Sub-saharan Africa |
| Search String | Coronavirus AND Diabetes AND Africa [*Title, Abstract & Keywords*) |
| Search limits/Filters | Publication years: 2020 - 2023 |
| Date of search | Initial: 10/05/2022  Updated: 22/03/2023 |
| Number of articles retrieved | 28 |
| Search notes | NA |

10. **Embase**

| Database access link | Via Ovid: <https://ovidsp.dc1.ovid.com/ovid-a/ovidweb.cgi?&S=OLLCFPLEBAACOLGGKPNJLGPKAHFAAA00&C=_main&tab=search&Main+Search+Page=1> |
| --- | --- |
| Search terms | Coronavirus  COVID-19  SARS-CoV-2  Diabetes  Diabetes mellitus  Africa |
| Search String | ((COVID-19 OR Coronavirus) AND Diabetes AND Africa).ab. |
| Search limits/Filters | Publication year: 2020 - 2023 |
| Date of search | Initial: 10/05/2022  Updated: 22/03/2023 |
| Number of articles retrieved | 113 |
| Search notes | NA |
