## Supplementary material for "Impact of COVID-19 on diabetes mellitus outcomes and care in sub-Saharan Africa: A scoping review": Themes

**Table S4: Major themes from qualitative, quantitative, and mixed methods studies as well as quantitative studies that reported open-ended results.**

| **Major theme** | **Sub-theme** | **Evidence summary** |
| --- | --- | --- |
| Patient-related health management challenges | Self-management challenges | Several studies (n=04) reported challenges with self-management of diabetes as reduced meal frequency, reduced physical activity and poor glycaemic control   - Reduced meal frequency   - *42.0% of patients reported reduced meal frequency (Ephraim RKD et al., 2021)*   - *57.7% of patients reported a reduction in daily meal frequency (Habineza JC et al., 2021)* - Reduced physical activity   - *43.1% reported reduced physical activity (Habineza JC et al., 2021)* - Poor glycaemic control among COVID-19 patients - *Uncontrolled diabetes (HbA1c>8%) was reported to be as high as 73.2% among people with diabetes infected with COVID-19 (Mash RJ et al., 2021). 78.6% of these patients were from rural areas.* - *86.5% of patients had HbA1c above 7% (median (IQR), 10% (8-12%) (Van der Westhuizen JN, et al., 2021).* |
|  | Affordability challenges | Several studies (n=02) reported affordability issues due to an increase in the cost of medication amidst reduced individual and household income during the pandemic.   - Increased cost of medicines   - *high medicine costs due to shortages from disruption of supply chains (Tagoe et al., 2023)* - Reduced individual or household income   - *80.8% of young adults with diabetes reported a drop in family income (Habineza JC et al., 2021)* |
|  | Health service accessibility challenges | Studies (n=03) reported health service access challenges during the pandemic as increased waiting time, and decreased transport options, particularly during multiple lockdowns.   - Increased clinic waiting time   - *Patients crowd outpatient departments from early morning, hoping to be seen early so they can go to their workplaces. However, this is not always possible because doctors attend to inpatients before seeing outpatients, resulting in long clinic waits (Tagoe et al., 2023).* - Limited availability transport options to healthcare facilities   - *There was a 16.2% decrease in the use of motorised transport during the COVID-19 pandemic and an 81.8% increase in foot travel (Habineza JC et al., 2021)*   - *Home delivery of medication was perceived as the solution to service access travel challenges (David JN et al., 2022)* |
| Diabetes care service delivery challenges | Health workforce challenges | Studies (n=02) reported hesitancy of health workers to attend healthcare stations, limited number of diabetes specialists and increased workload as COVID-19 pandemic-related challenges.   - Health worker hesitance (Delobelle AP et al., 2022) - Limited number of diabetes specialists (Tagoe et al., 2023) - Increased workload on community health workers (Delobelle AP et al., 2022) |
|  | Healthcare infrastructure challenges | The study (n=01) reported the COVID-19 pandemic resulted in limited physical clinic space due to the overwhelming patient number   - Limited physical space to run diabetes clinic services (Tagoe et al., 2023) |
|  | Health information challenges | The study (n=01) showed that COVID-19 was associated with poor patient information and records management due to work overload and fear of infection.   - Poor patient information and records management (Brey Z et al., 2020) |
|  | Medicines and medical supplies | The study (n=01) reported worsened shortages in supplies of medicines and medical supplies due to disruptions in supply chains.   - Shortage of medicine and medical supplies as a result of supply chain disruptions (Tagoe et al., 2023) |
| Re-organisation of diabetes care delivery | Patient-level reorganisation of care access | Studies (n=03) reported home delivery of medicines to patients as an intervention in response to COVID-19 pandemic challenges of patient accessibility, risk of infection and fear of travel.   - Home delivery of medication to reduce infection risk, and combat fear of infection among people with diabetes while mitigating the health system access challenges faced by patients due to shutdown of transport facilities during multiple lockdowns.   - Home delivery of patient medical supplies was introduced (Brey Z et al., 2020; Delobelle AP et al., 2022; David JN et al., 2022) |
|  | Clinic-level reorganisation of management | The study reported the cancellation of open walk-in non-communicable disease clinic days and the creation of new methods of booking systems to manage the number of patients visiting the clinic at a particular time to avoid overcrowding.   - Cancellation of the routine noncommunicable disease clinics (Delobelle AP et al., 2022) - Institution of clinic booking system to manage patient appointments (Delobelle AP et al., 2022) |
|  | Community-level re-organisation of community health worker services | Studies (n=03) reported that community health workers were empowered to provide community monitoring and follow-up of chronic diseases, including diabetes.   - Engagement of community health workers (Brey Z et al., 2020; Delobelle AP et al., 2022; David JN et al., 2022) |
